# Patient-reported symptom profiles in vitamin B_12_ deficiency and pernicious anemia: A cross-sectional study using latent class analysis

**DOI:** 10.64898/2026.06.15.26355707

**Authors:** Brooke M. Morris, Alfie Thain, Blair Coe Schweiger, Ebba Nexø, Andrew McCaddon, Bruce H. R. Wolffenbuttel, Ralph Green, David H. Alpers, Marie Joe Dib, Petra Visser, Katrina Burchell, Kourosh R. Ahmadi, Austin W. Reynolds

## Abstract

**Background:** Pernicious anemia is a chronic autoimmune disease characterized by impaired of vitamin B_12_ absorption and deficiency. Current diagnostic approaches rely on laboratory biomarkers with limitations in sensitivity and specificity, leading to diagnostic uncertainty and potentially delayed and/or suboptimal treatment. The 2024 NICE guidelines acknowledge that treatment frequency should be guided by individual symptom response rather than current one-size-fits-all schedules; however, the symptom heterogeneity underpinning this recommendation remains poorly characterized.

**Methods and Findings:** We conducted secondary analysis of symptom survey data from 1,117 members of the Pernicious Anaemia Society (PAS) collected between August 2010 and November 2012. We applied latent class analysis (LCA) to 46 self-reported indicators comprising demographic characteristics, symptoms, and comorbid conditions, to identify distinct symptom-based subtypes and assess whether such subgroups can inform more tailored management of PA. Associations between symptom subtypes and diagnostic test results, age when symptoms first started, symptom duration, and treatment satisfaction were examined using chi-square and Fisher’s exact tests.

The best model included three distinct symptom subtypes: High Burden (n=334, 29·9%), Moderate Burden (n=613, 54·9%), and Low Burden (n=170, 15·2%). The High Burden subtype exhibited fatigue (99·4%), cognitive dysfunction (97·9% memory loss), neurological manifestations (95·5% clumsiness), and suicidal thoughts (41·6%). Intrinsic factor antibody (IFA) status did not differ significantly across subtypes (χ^2^=1·04, df=2, p=0·593). Age at symptom onset differed significantly across subtypes (p=0·002), with the Low Burden subtype overrepresented in older age groups. The High Burden subtype had the longest diagnostic delays (56·9% >2 years). Standard three-monthly injections showed low satisfaction across all classes (High Burden: 8·5%, Moderate Burden: 23·7%, Low Burden: 30·7%).

**Conclusions:** Symptom-based stratification identifies clinically meaningful subgroups independent of IFA status, supporting tailored, symptom-guided treatment rather than the current piecemeal approach. The inverse association between high symptom burden and age of onset warrants clinical attention. The co-occurrence of markedly elevated rates of depression and suicidal thoughts in the High Burden subtype suggest that integrated mental health assessments should form part of routine clinical management.

## Introduction

Vitamin B_12_ is an essential micronutrient obtained almost exclusively from animal products. It plays critical roles in numerous physiological processes including myelination, DNA synthesis, methyltransferase activity and gene expression regulation.[1,2] Deficiency is surprisingly common, affecting approximately 6% of people under age 60 and 10-20% of the elderly population.[3,4] B_12_ deficiency can result from various factors including malabsorption disorders, medication effects (proton-pump inhibitors and metformin), and vegetarian or vegan diets.[5,6] A particularly concerning cause is autoimmune destruction of an essential component for the normal B_12_ absorption pathway, leading to pernicious anemia (PA).[7]

PA represents the end-stage of autoimmune gastritis (AIG), a complex and heterogeneous condition in which antibodies target gastric parietal cells in the stomach fundus and corpus.[8–10] These cells produce intrinsic factor, a protein essential for B_12_ absorption in the small intestine.[11] Without treatment, the resulting B_12_ deficiency progressively leads to neurological dysfunction, cognitive decline, severely compromised quality of life (QoL), and megaloblastic anemia.[12]

Although PA is commonly considered rare, affecting only 0·1% of the general population and 2-3% of older adults, diagnostic challenges lead to significant underestimation of its true prevalence.[13,14] Distinguishing PA from other causes of B_12_ deficiency presents a fundamental clinical challenge, as low B_12_, high methylmalonic acid (MMA) or homocysteine cannot differentiate autoimmune-mediated PA from dietary insufficiency, malabsorption due to medications, or other gastrointestinal conditions. Current recommendations for diagnostic approaches distinguishing PA from other causes of B_12_ deficiency rely on measures of intrinsic factor antibodies (IFA) sometimes including parietal cell antibodies (PCA) and occasionally endoscopy with biopsy to assess atrophic gastritis.[15–19] However, these tests have limitations in both accuracy and clinical application. IFA show high specificity (95-100%) for AIG and PA but suffer from poor sensitivity (approximately 27%).[20] PCA offer higher sensitivity (81%) but reduced specificity (90%). Combining both antibody tests improves overall diagnostic accuracy to 60·6% sensitivity with maintained 95-100% specificity, reaching up to 73% sensitivity in patients with clinical PA.[20] Added to this, many General Practitioners in the United Kingdom (UK) have not adopted these extensive testing approaches.[21] The inconsistent diagnostic outcomes are compounded by antibody testing’s variable performance at different disease stages.[13,15,20,22,23]

Critically, patients with and without definitive PA confirmation face substantial barriers to accessing adequate treatment. The Pernicious Anaemia Society (PAS), a UK-based patient advocacy group, conducted two surveys (2012 and 2022) aimed at better understanding patient experiences and needs.[24,25] Previous analysis of diagnostic testing in PAS membership highlights diagnostic uncertainty, which directly impacts treatment access and QoL[24]. Recent evidence also shows that patients whose injections were suspended or switched to oral supplementation during the COVID-19 pandemic reported worsening symptoms and reduced well-being compared to those who maintained their usual injection regimens, or those that were able to optimize their treatment frequency.[26] Furthermore, a cross-sectional study found that 4 out of 10 patients with B_12_ deficiency reported self-injecting B_12_ obtained outside the National Health Service (NHS), primarily due to difficulty accessing treatment in primary care.[27] A review from the UK’s National Institute for Health and Care Excellence (NICE) acknowledges this treatment gap, noting that intramuscular B_12_ is generally reserved for confirmed malabsorption cases, creating uncertainty when IFA are negative despite clinical B_12_ deficiency.[28] This gatekeeping based on serological results rather than symptom burden means that treatment access is determined by diagnostic classification rather than clinical need, perpetuating a one-size-fits-all approach that may leave many patients undertreated.[24]

We propose that clinically relevant symptoms may be more important than diagnostic labels, and that treatment should be guided by symptom control rather than diagnostic category. In line with this, the 2012 PAS survey, the dataset analyzed in the present study, found that while some patients respond well to the standard UK regimen, >60% of PAS members are dissatisfied with their treatment frequency and have trouble in maintaining an acceptable QoL. Traditional diagnostic categories might not adequately capture these differences, in response to standardized treatment, possibly explaining why they fail to satisfy many patients’ needs. Updated NICE guidelines acknowledge this by recommending that patients receive intramuscular hydroxocobalamin injections titrated to symptom severity or response.[28–30] However, this recommendation is qualified by the requirement to prescribe within the medication’s licensed dosing schedule, which in practice may lead clinicians to default to standardized two- to three-monthly intervals regardless of individual symptom burden. Improving the quality of care for chronic, life-long conditions like PA therefore requires systematic changes guided by research advances and frameworks such as the chronic care model (CCM).[31] This advocates for delivery system restructure within clinical information systems to track patient outcomes, and self-management support to empower patients as partners in their care.[31] Identifying symptom subtypes within this heterogeneous population is a critical step toward enabling such individualized approaches. This is particularly relevant given that the 2024 NICE guidelines, while recommending symptom-titrated treatment, describe symptoms in vague terms and provide limited guidance for patients who presenting with B_12_ deficiency without definitive PA confirmation.

Addressing both the NICE implementation gap and the broader challenge of tailoring treatment to diagnostic uncertainty requires first identifying which patient subgroups exist and what their differential needs are; the foundation of the CCM-informed, personalized care. In this study, we applied latent class analysis (LCA), a person-centered statistical approach that identifies unobserved subgroups within populations, to examine data from the 2012 PAS patient survey. Unlike previous descriptive studies, this approach identifies distinct symptom patterns and assesses their agreement with current diagnostic categories.[25,32] We examined relationships between symptom profiles, demographic characteristics, pre-diagnosis symptom duration, and treatment satisfaction across patients with B_12_-related disorders, independent of formal diagnostic classification, to evaluate if symptom burden provides greater guidance for treatment decisions compared to diagnostic status. Our primary hypothesis is that patients experiencing B_12_-related disorders comprise distinct subgroups characterized by their symptom presentations, and that these subgroups extend beyond the traditional diagnostic boundary separating PA from B_12_ deficiency of uncertain etiology. Our secondary hypothesis is that these symptom-defined subgroups reveal differential treatment responses and satisfaction levels, with symptom severity and pattern serving as stronger predictors of outcomes than a clinical diagnosis.

## Methods

### Study design

In this cross-sectional retrospective secondary analysis, we applied latent class analysis (LCA) to survey data collected by the PAS from their patient cohort (**S1 Table**).[24] This study follows the Strengthening the Reporting of Observational Studies in Epidemiology (STROBE) guidelines for cross-sectional studies.[33] The initial study was conducted in the UK through the PAS and was designed to evaluate current care without randomization or intervention allocation.[34]

### Participants

All PAS members (n=3700) were deemed eligible, and no exclusion criteria were applied during the data collection stage. The survey was made available online through the PAS website (https://pernicious-anemia-society.org/), and paper copies were mailed to members without Internet access.

### Procedures

Data were collected over a 15-month period between August 2010 and November 2012 using a comprehensive four-section survey instrument. The survey collected: (a) demographic factors such as sex (male, female), age at diagnosis, and family history of PA; (b) symptoms experienced prior to diagnosis across seven physiological domains: general, neurological, respiratory, cardiovascular, gastrointestinal, genitourinary, and emotional; (c) coexisting medical conditions across eight areas: general, neurological, dermatologic, cardiovascular, musculoskeletal, endocrine, gastrointestinal, and emotional; and (d) diagnostics including biomarkers and endoscopy results, treatment type and frequency, and satisfaction with treatment. All symptom and disease variables were binary (presence/absence). Participants were specifically asked to report symptoms experienced prior to diagnosis rather than current symptoms, which helped to focus this analysis on symptom descriptions for use in a diagnostic and treatment management model.

### Statistical analysis

A missing data pattern analysis was conducted to assess potential non-response bias; complete data were available for all 46 LCA indicator variables, with missing data observed only in post-hoc outcome variables (**S1 Fig**). To assess potential recall bias associated with retrospective symptom reporting, Spearman rank correlations were used to examine the relationship between duration of symptoms before diagnosis and individual symptom endorsement across all 35 symptom indicators.[35]

### Latent Class Analysis

We employed LCA to determine distinct symptom profiles within the study population. LCA is a person-centered statistical approach that identifies unobserved subgroups (latent classes) within a population based on response patterns across multiple observed variables.[36] We selected symptoms and comorbid conditions as observed indicators for the LCA based on prevalence within our sample. Following the methodological recommendations of Sinha et al. (2021), we included only indicators with a prevalence of at least 10%.[37] This threshold helps prevent the identification of latent classes that are exclusively based on rare indicators, which result in the classes driven by rare responses rather than meaningful population heterogeneity. This resulted in 46 indicators comprising three demographic indicators, 35 symptoms, and eight comorbid condition indicators representing the most common manifestations of B_12_ deficiency in our population.

Models with 2-10 classes were estimated and compared using multiple fit indices including Akaike Information Criterion (AIC), Bayesian Information Criterion (BIC), and sample-size adjusted BIC (aBIC), along with entropy and Lo-Mendell-Rubin adjusted likelihood ratio tests (LMR-LRT). Lower information criterion values (AIC, BIC, aBIC) indicate better model fit, with BIC typically given greater weight as it penalizes model complexity more strongly than AIC and aBIC, reducing the risk of overfitting. Higher entropy values (closer to 1·0) indicate better classification quality, and a significant LMR-LRT p-value indicates that the k-class model fits significantly better than the (k-1)-class model. Model selection involved visual examination of elbow plots for all information criteria to identify where diminishing returns in fit improvement occurred.

### Post-hoc analysis

To examine the relationship between symptom burden and autoimmune etiology, IFA test results were analyzed across the three latent class subtypes. Only participants with a reported IFA result (either positive or negative) were included in the analysis (n=222, 19·9% of the analytic sample). A chi-square test was used to examine whether IFA positivity differed significantly across the three symptom burden subtypes.

Associations between latent class membership and two patient characteristics were examined. Duration of symptoms before receiving a diagnosis was collected across 11 ordered categories: 1-week, 2-weeks, 1-month, 2-months, 3-months, 6-months, 1-year, 2-years, 5-years, 10-years, and 10+ years. These timeframes were selected based on their clinical relevance. Chi-square analysis with Cramér’s V effect size tested whether symptom duration differed across symptom burden subtypes. Age when symptoms first started was analyzed as a categorical variable in nine ranges (0-10, 11-20, 21-30, 31-40, 41-50, 51-60, 61-70, 71-80, 81-90) and evaluated using chi-square analysis to test for associations with symptom burden subtype. To identify which specific age and subtype combinations drove any observed associations, standardized residuals were calculated; absolute values above 1·96, corresponding to p<0·05 under the standard normal distribution, were treated as statistically significant. Participants with missing data for age when symptoms commenced were excluded listwise from this analysis (n=1,013, 90·7% of the analytic sample).

Treatment frequency and satisfaction rates were examined across symptom burden subtypes. Frequency categories were based on standard and self-directed administration schedules for B_12_ replacement therapy and collapsed where appropriate: weekly or more (including daily, more than once a day, and weekly), monthly, two-monthly, three-monthly, other, or no treatment. Participants with missing treatment frequency data (n=155, 13·9% of analytic sample) or missing satisfaction responses (n=37) were excluded listwise. Fisher’s exact tests with 10,000 Monte Carlo simulations compared satisfaction rates for each frequency category against three-monthly injections as the reference category. Free-text responses in the “Other” category were categorized into five groups based on injection interval: loading phase, very frequent (every two weeks or less), extended standard (3-11 weeks), extended interval (12 weeks or more), and no treatment or pre-treatment.

All analyses were conducted using Mplus Version 8 for LCA and R Statistical Software (version 4·0·3; R Core Team 2021) for descriptive statistics and post-hoc analyses.[38,39]

### Role of the funding source

The funding source did not provide financial support for this research and was not involved in study design, data collection, data analysis, interpretation of results, or manuscript preparation. The authors maintained complete independence in all aspects of the research process and retain full responsibility for the content of this manuscript.

## Results

Of the 3,700 PAS members invited to participate, 1,184 completed the survey (response rate, 32·0%). Four respondents were excluded from analysis: one did not answer any questions, two did not provide sex information, and one answered only three questions, yielding a working sample of 1,180. A further 63 participants were excluded; they reported neither a formal diagnosis of B_12_ deficiency or PA nor biomarker evidence of B_12_ deficiency (low serum B_12_, positive IFA, or abnormal gastroscopy). This resulted in a final analytic sample of 1,117 participants (**Fig 1**). The analytic included PAS members across the diagnostic spectrum, encompassing those who reported a formal PA diagnosis and B_12_ deficiency without PA diagnosis.

**Fig 1.**
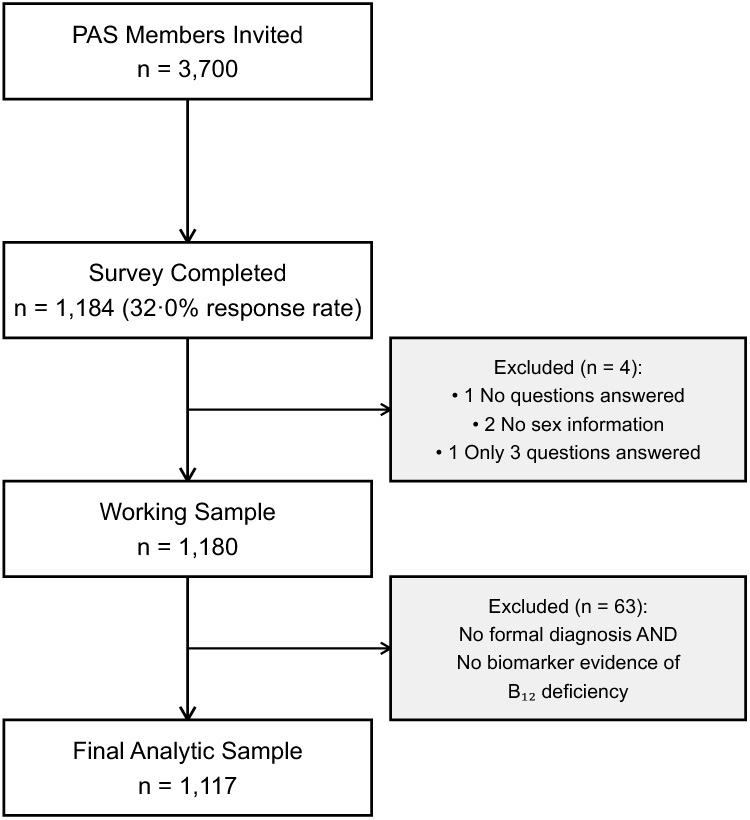
Study Flow Diagram. The flow chart illustrates participant recruitment and inclusion. From the 3,700 Pernicious Anaemia Society (PAS) members invited, 1,184 completed the survey (32·0% response rate). Four participants were excluded due to insufficient data (one provided no responses, two did not provide sex information, and one answered only three questions), yielding a working sample of 1,180. A further 63 participants were excluded who reported they had neither a formal diagnosis of B12 deficiency or pernicious anemia nor reported biomarker evidence of B12 deficiency (low serum B12, positive intrinsic factor antibody, or abnormal gastroscopy), resulting in a final analytic sample of N=1,117.

Our LCA comprised data on 1,117 participants from the PAS, with age when symptoms first started ranging from 0-90 years. Most participants were female (n=913, 81·6%) and a majority of participants reported a formal diagnosis of PA (n=853, 76·4%), while 243 (21·7%) reported B_12_ deficiency without PA diagnosis. A further 13 (1·2%) reported no formal diagnosis but were retained based on reported biomarker evidence of B_12_ deficiency. Complete data were available for all 46 LCA indicator variables across the full analytic sample. Missing data were only observed in post-hoc outcome variables: age when symptoms first started (9·3%), duration of symptoms before diagnosis (6·4%), and treatment satisfaction (8·1%; **S1 Fig**). While 33 of 35 symptoms (94·3%) showed statistically significant correlations with symptom duration, the magnitude of these correlations was weak (ranging from -0·032 to 0·205), suggesting minimal impact of recall bias on primary findings (**S2 Fig, S2 Table)**.[35] All symptom and comorbidity data represent self-reported patient experience and should be interpreted as reflecting perceived symptom burden rather than clinically verified diagnoses.

We tested latent class models with 2-10 classes to identify distinct symptom subtypes among participants with B_12_ deficiency and/or PA (**Table 1**). The 3-class solution emerged as the best fit and most parsimonious based on convergence of fit indices, LMR-LRT significance, and adequate class sizes. This model showed significant improvement over the 2-class solution across all information criteria, with lower values for AIC (55866 vs. 56733), BIC (56568 vs. 57200), and sample-size adjusted BIC (aBIC: 56123 vs. 56904) (**S3 Table, S3 Fig**). While AIC and aBIC continued to decrease with additional classes, BIC showed an inflection point at the 3-class solution before increases became more gradual, suggesting diminishing returns in model fit beyond this point. The 3-class model fit was significantly better than the 2-class solution (p<0·0001) as confirmed with the LMR-LRT. Importantly, the LMR-LRT test for the 4-class solution was non-significant (p=0·51), indicating no statistically meaningful improvement over the 3-class model despite the slightly lower BIC. Additionally, the 3-class solution showed high entropy (0·86, indicating clear separation between classes and robust classification of participants. The three identified subtypes are described in detail below, labelled High Burden, Moderate Burden, and Low Burden based on cumulative symptom prevalence across domains.

**Table 1.**
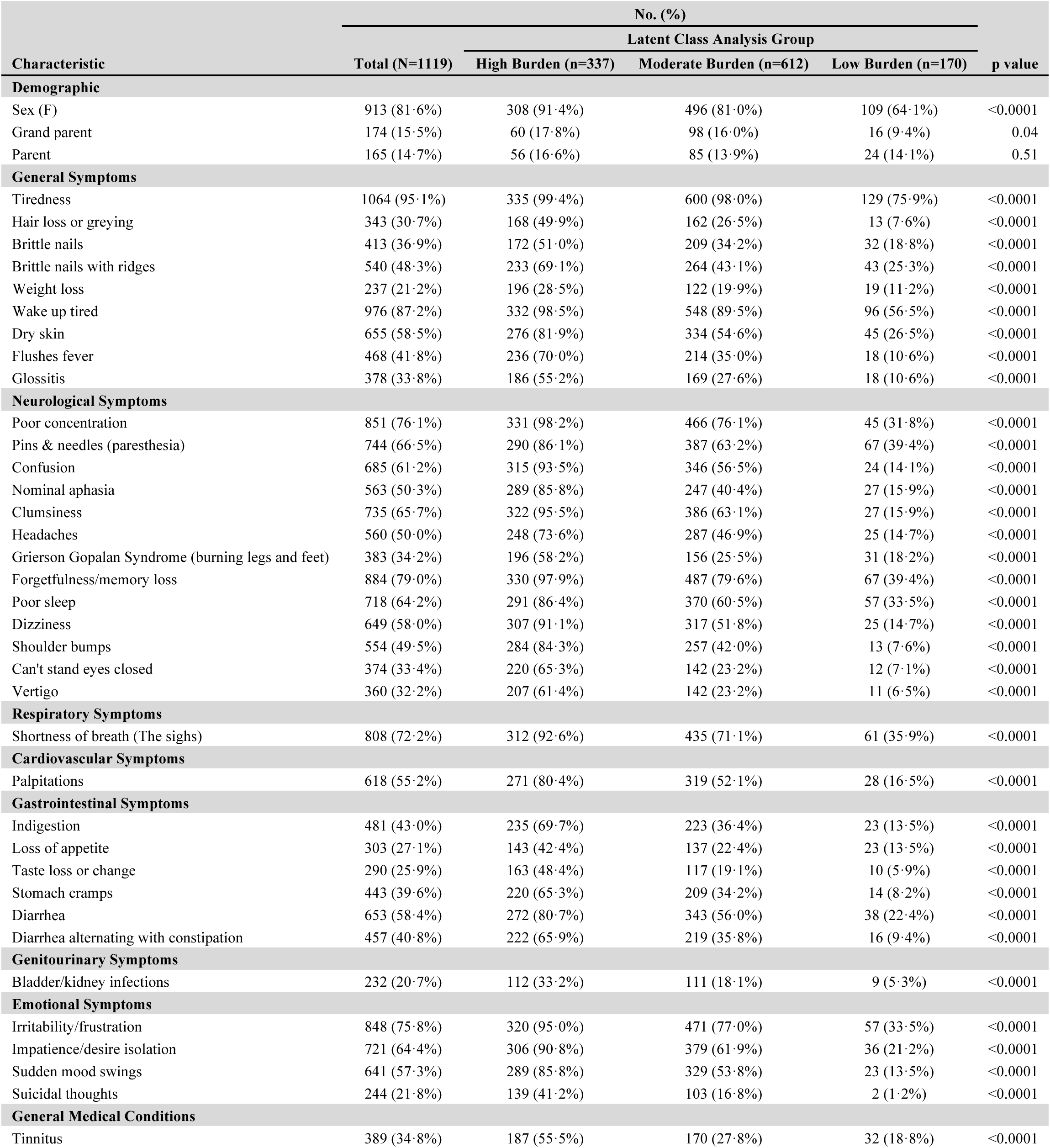
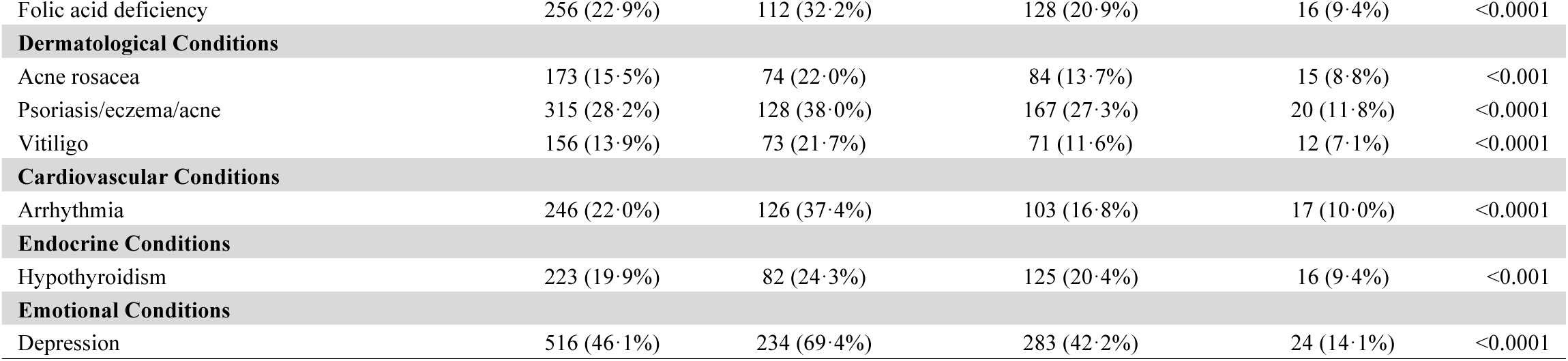
Demographic characteristics, symptoms, and comorbidities across latent classes of vitamin B12 deficiency (N=1119) This table presents the distribution of demographic characteristics, clinical symptoms, and comorbid conditions across the three latent classes identified through latent class analysis. Class 3 (High Burden, n=337) demonstrates the highest symptom prevalence across all domains, Class 2 (Moderate Burden, n=612) exhibits intermediate symptom prevalence, and Class 1 (Low Burden, n=170) shows substantially lower symptom prevalence. Values represent number (percentage) of participants within each class reporting the presence of each indicator. P-values derive from chi-square tests comparing prevalence across classes. Notably, 45 of 46 indicators (97·8%) showed statistically significant differences across classes (p<0·05), with only parental history of pernicious anemia (p=0·51) showing similar distribution across all three groups.

| Characteristic | No. (%) |  |  |  | p value |
| --- | --- | --- | --- | --- | --- |
|  | Total (N=1119) | Latent Class Analysis Group |  |  |  |
|  |  | High Burden (n=337) | Moderate Burden (n=612) | Low Burden (n=170) |  |
| Demographic |  |  |  |  |  |
| Sex (F) | 913 (81.6%) | 308 (91.4%) | 496 (81.0%) | 109 (64.1%) | <0.0001 |
| Grand parent | 174 (15.5%) | 60 (17.8%) | 98 (16.0%) | 16 (9.4%) | 0.04 |
| Parent | 165 (14.7%) | 56 (16.6%) | 85 (13.9%) | 24 (14.1%) | 0.51 |
| General Symptoms |  |  |  |  |  |
| Tiredness | 1064 (95.1%) | 335 (99.4%) | 600 (98.0%) | 129 (75.9%) | <0.0001 |
| Hair loss or greying | 343 (30.7%) | 168 (49.9%) | 162 (26.5%) | 13 (7.6%) | <0.0001 |
| Brittle nails | 413 (36.9%) | 172 (51.0%) | 209 (34.2%) | 32 (18.8%) | <0.0001 |
| Brittle nails with ridges | 540 (48.3%) | 233 (69.1%) | 264 (43.1%) | 43 (25.3%) | <0.0001 |
| Weight loss | 237 (21.2%) | 196 (28.5%) | 122 (19.9%) | 19 (11.2%) | <0.0001 |
| Wake up tired | 976 (87.2%) | 332 (98.5%) | 548 (89.5%) | 96 (56.5%) | <0.0001 |
| Dry skin | 655 (58.5%) | 276 (81.9%) | 334 (54.6%) | 45 (26.5%) | <0.0001 |
| Flushes fever | 468 (41.8%) | 236 (70.0%) | 214 (35.0%) | 18 (10.6%) | <0.0001 |
| Glossitis | 378 (33.8%) | 186 (55.2%) | 169 (27.6%) | 18 (10.6%) | <0.0001 |
| Neurological Symptoms |  |  |  |  |  |
| Poor concentration | 851 (76.1%) | 331 (98.2%) | 466 (76.1%) | 45 (31.8%) | <0.0001 |
| Pins & needles (paresthesia) | 744 (66.5%) | 290 (86.1%) | 387 (63.2%) | 67 (39.4%) | <0.0001 |
| Confusion | 685 (61.2%) | 315 (93.5%) | 346 (56.5%) | 24 (14.1%) | <0.0001 |
| Nominal aphasia | 563 (50.3%) | 289 (85.8%) | 247 (40.4%) | 27 (15.9%) | <0.0001 |
| Clumsiness | 735 (65.7%) | 322 (95.5%) | 386 (63.1%) | 27 (15.9%) | <0.0001 |
| Headaches | 560 (50.0%) | 248 (73.6%) | 287 (46.9%) | 25 (14.7%) | <0.0001 |
| Grierson Gopalan Syndrome (burning legs and feet) | 383 (34.2%) | 196 (58.2%) | 156 (25.5%) | 31 (18.2%) | <0.0001 |
| Forgetfulness/memory loss | 884 (79.0%) | 330 (97.9%) | 487 (79.6%) | 67 (39.4%) | <0.0001 |
| Poor sleep | 718 (64.2%) | 291 (86.4%) | 370 (60.5%) | 57 (33.5%) | <0.0001 |
| Dizziness | 649 (58.0%) | 307 (91.1%) | 317 (51.8%) | 25 (14.7%) | <0.0001 |
| Shoulder bumps | 554 (49.5%) | 284 (84.3%) | 257 (42.0%) | 13 (7.6%) | <0.0001 |
| Can't stand eyes closed | 374 (33.4%) | 220 (65.3%) | 142 (23.2%) | 12 (7.1%) | <0.0001 |
| Vertigo | 360 (32.2%) | 207 (61.4%) | 142 (23.2%) | 11 (6.5%) | <0.0001 |
| Respiratory Symptoms |  |  |  |  |  |
| Shortness of breath (The sighs) | 808 (72.2%) | 312 (92.6%) | 435 (71.1%) | 61 (35.9%) | <0.0001 |
| Cardiovascular Symptoms |  |  |  |  |  |
| Palpitations | 618 (55.2%) | 271 (80.4%) | 319 (52.1%) | 28 (16.5%) | <0.0001 |
| Gastrointestinal Symptoms |  |  |  |  |  |
| Indigestion | 481 (43.0%) | 235 (69.7%) | 223 (36.4%) | 23 (13.5%) | <0.0001 |
| Loss of appetite | 303 (27.1%) | 143 (42.4%) | 137 (22.4%) | 23 (13.5%) | <0.0001 |
| Taste loss or change | 290 (25.9%) | 163 (48.4%) | 117 (19.1%) | 10 (5.9%) | <0.0001 |
| Stomach cramps | 443 (39.6%) | 220 (65.3%) | 209 (34.2%) | 14 (8.2%) | <0.0001 |
| Diarrhea | 653 (58.4%) | 272 (80.7%) | 343 (56.0%) | 38 (22.4%) | <0.0001 |
| Diarrhea alternating with constipation | 457 (40.8%) | 222 (65.9%) | 219 (35.8%) | 16 (9.4%) | <0.0001 |
| Genitourinary Symptoms |  |  |  |  |  |
| Bladder/kidney infections | 232 (20.7%) | 112 (33.2%) | 111 (18.1%) | 9 (5.3%) | <0.0001 |
| Emotional Symptoms |  |  |  |  |  |
| Irritability/frustration | 848 (75.8%) | 320 (95.0%) | 471 (77.0%) | 57 (33.5%) | <0.0001 |
| Impatience/desire isolation | 721 (64.4%) | 306 (90.8%) | 379 (61.9%) | 36 (21.2%) | <0.0001 |
| Sudden mood swings | 641 (57.3%) | 289 (85.8%) | 329 (53.8%) | 23 (13.5%) | <0.0001 |
| Suicidal thoughts | 244 (21.8%) | 139 (41.2%) | 103 (16.8%) | 2 (1.2%) | <0.0001 |
| General Medical Conditions |  |  |  |  |  |
| Tinnitus | 389 (34.8%) | 187 (55.5%) | 170 (27.8%) | 32 (18.8%) | <0.0001 |
| Folic acid deficiency | 256 (22·9%) | 112 (32·2%) | 128 (20·9%) | 16 (9·4%) | <0.0001 |
| **Dermatological Conditions** |  |  |  |  |  |
| Acne rosacea | 173 (15·5%) | 74 (22·0%) | 84 (13·7%) | 15 (8·8%) | <0.001 |
| Psoriasis/eczema/acne | 315 (28·2%) | 128 (38·0%) | 167 (27·3%) | 20 (11·8%) | <0.0001 |
| Vitiligo | 156 (13·9%) | 73 (21·7%) | 71 (11·6%) | 12 (7·1%) | <0.0001 |
| **Cardiovascular Conditions** |  |  |  |  |  |
| Arrhythmia | 246 (22·0%) | 126 (37·4%) | 103 (16·8%) | 17 (10·0%) | <0.0001 |
| **Endocrine Conditions** |  |  |  |  |  |
| Hypothyroidism | 223 (19·9%) | 82 (24·3%) | 125 (20·4%) | 16 (9·4%) | <0.001 |
| **Emotional Conditions** |  |  |  |  |  |
| Depression | 516 (46·1%) | 234 (69·4%) | 283 (42·2%) | 24 (14·1%) | <0.0001 |

The High Burden subtype (n=334, 30·0%) was characterized by the highest symptom burden across all domains, exhibiting fatigue (99·4%) and waking up tired (98·5%), accompanied by cognitive dysfunction, including poor concentration (98·5%), memory loss (97·9%), and confusion (93·7%). Neurological manifestations were prominent, with high rates of clumsiness (95·5%), paresthesia (86·2%), and dizziness (91·0%), together with significant mood disturbances, including irritability (94·9%), and impatience (90·7%). Notably, this subtype had the highest rate of suicidal thoughts (41·6%) and highest prevalence of affected grandparents (18·0%). Comorbid conditions were common across multiple systems, including cardiovascular (arrhythmia 37·4%), endocrine (hypothyroidism 24·3%), dermatological conditions (vitiligo 21·6%; psoriasis/eczema/acne 38·0%), and general medical conditions (tinnitus 55·4%; folic acid deficiency 32·2%). Depression, classified as a distinct emotional condition rather than a symptom, was notably prevalent in this subtype (69·8%).

The Moderate Burden subtype (n=613, 54·9%) presented with intermediate symptom prevalence. Fatigue in this subtype remained high (98·0%), but cognitive symptoms were less prevalent, with lower rates of memory loss (79·6%), poor concentration (76·2%), and confusion (56·6%). Neurological manifestations followed a similar pattern, with moderate rates of paresthesia (63·3%) and clumsiness (63·3%). Mood disturbances were present but less pronounced than the High Burden subtype, including suicidal thoughts in 16·8%. Depression was reported by 42·3% of this subtype, reflecting a similar distinct condition pattern as observed in the High Burden class.

The Low Burden subtype (n=170, 15·2%) exhibited a markedly reduced symptom prevalence and showed a lower female predominance (64·1% compared to 81·0% and 91·4% for Moderate and High Burden subtypes) than the other subtypes. Fatigue, while still present (76·5%), was significantly less prevalent than in other subtypes. Cognitive dysfunction was minimal, with lower rates of memory loss (40·6%), and confusion (14·7%). Neurological symptoms were rare, with minimal clumsiness (15·9%) and dizziness (14·7%). This subtype had minimal mental health impact, with depression affecting only 14·1% and suicidal thoughts in just 1·2%. This subtype represents a substantially milder clinical phenotype with more limited multisystem involvement compared to the High Burden and Moderate Burden subtypes.

IFA test results were available for 222 participants (19·9% of the analytic sample), limiting the interpretability of antibody-based comparisons across subtypes. Among those tested, positive IFA results were similarly distributed across subtypes: 49·4% in the High Burden subtype, 56·2% in the Moderate Burden subtype, and 57·1% in the Low Burden subtype (χ²=1·04, df=2, p=0·593; **Table 2).** These findings suggest that IFA positivity does not differentiate between symptom burden subtypes.

**Table 2.** Intrinsic factor antibody (IFA) test results across latent classes (n=222 tested, 19·8% of analytic sample). The proportion of positive IFA results is reported descriptively for each symptom burden subtype. A chi-square test comparing IFA positivity across the three subtypes showed no significant difference (χ^2^=1·08, df=2, p=0·584), indicating that the likelihood of a positive IFA result was similar regardless of symptom burden subtype. These findings suggest that autoimmunity etiology, as measured by IFA, does not differentiate symptom burden subtypes. Interpretation should be made with caution given the poor sensitivity of IFA (approximately 27%); a negative result does not exclude pernicious anemia.^20^

| Class Subtype | N (%) |  |  |  | p value |
| --- | --- | --- | --- | --- | --- |
|  | IFA Positive | IFA Negative | Total Tested | % Positive |  |
| High Burden | 45 | 46 | 91 | 49.5% | 1.00 |
| Moderate Burden | 58 | 45 | 103 | 56.3% | 0.24 |
| Low Burden | 16 | 12 | 28 | 57.1% | 0.57 |

Age at symptom onset differed significantly across subtypes (χ2=37・07, df=16, p=0・002, Cramer’s V=0・14), with the Low Burden subtype showing overrepresentation in older age groups, particularly ages 71-80 (z=3·54) and 61-70 (z=2·52). The High Burden subtype showed overrepresentation in the 31-40 age group (z=2·04) and underrepresentation in the 71-80 age group (z=-2·50) suggesting that higher symptom burden is associated with younger age of symptom onset. The Moderate Burden subtype demonstrated a more balanced age distribution closely matching the overall sample (**S4 Fig, S4 Table**; n=1,013).

Symptom duration pre-diagnosis varied significantly between subtypes (χ²=115·47, df=4, p<0·001, Cramér’s V=0·23; n=1,046). The High Burden subtype experienced longer pre-diagnosis symptom periods, with 56·9% reporting symptoms of more than 2 years before diagnosis, compared with 40·6% in the Moderate Burden subtype and dropping to 19·2% in the Low Burden subtype. Only 2·5% of the High Burden subtype received a diagnosis within six months of symptom onset, compared to 4·4% in the Moderate Burden subtype and 21·8% in the Low Burden subtype (**S5 Fig, S6 Fig**).

Three-monthly injections showed consistently low satisfaction across all subtypes (High Burden: 8·5%; Moderate Burden: 23·7%; Low Burden: 30·7%). In the High Burden and Moderate Burden subtypes, all frequencies except two-monthly were associated with significantly higher satisfaction than three-monthly injections (all p<0·001). Two-monthly injections approached but did not reach significance in the High Burden subtype (p=0·063), while all frequencies including the two-monthly reached significance in the Moderate Burden subtype (p<0·001). The Low Burden subtype showed significance in the monthly (p=0·037) and “Other” (p=0·002), suggesting greater acceptability of less frequent treatment in this group. The “Other” category showed the highest satisfaction rates across all subtypes (High Burden: 45·9%; Moderate Burden: 58·8%; Low Burden: 73·3%; **Table 3**, **S5 Table)**.

**Table 3.**
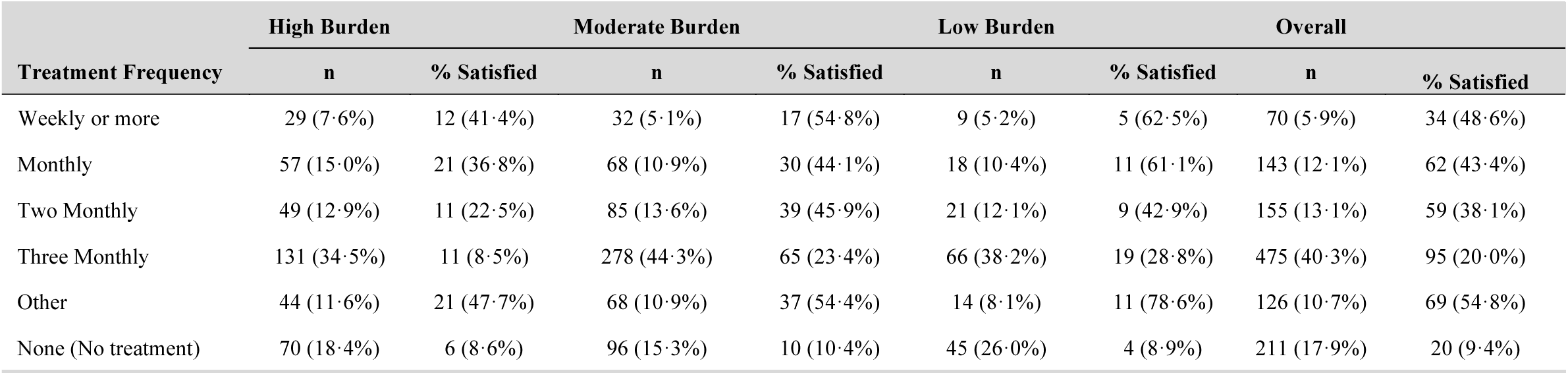
Treatment Frequency and Satisfaction Rates Across Latent Classes. Distribution of B12 replacement therapy frequency and corresponding satisfaction rates within each symptom-based latent class. Values represent number of participants (percentage within class) receiving each treatment frequency, with satisfaction rates shown as number satisfied/total number (percentage satisfied) for each frequency category. Statistical tests confirmed highly significant associations between treatment frequency and satisfaction within all classes (High Burden: χ2=41・15, df=4, p<0・001; Moderate Burden: χ2=38・51, df=4, p<0・001; Low Burden: Fisher’s exact test p= 0・004), with None treatment and missing satisfaction responses excluded from tests. Notable findings include consistently poor satisfaction with three-monthly injections across all classes (High Burden: 8・5%, Moderate Burden: 23・4%, Low Burden: 28・8%), while optimal treatment frequencies varied by class: Weekly or more administration for High Burden class, weekly for Moderate Burden class, and monthly for Low Burden class. The “Other” category (see **S5 Table** for details), representing alternative treatment approaches, showed high satisfaction rates across all classes (47・7-78・6%), suggesting patients benefit from personalized protocols beyond standard administration schedules.

| Treatment Frequency | High Burden |  | Moderate Burden |  | Low Burden |  | Overall |  |
| --- | --- | --- | --- | --- | --- | --- | --- | --- |
|  | n | % Satisfied | n | % Satisfied | n | % Satisfied | n | % Satisfied |
| Weekly or more | 29 (7.6%) | 12 (41.4%) | 32 (5.1%) | 17 (54.8%) | 9 (5.2%) | 5 (62.5%) | 70 (5.9%) | 34 (48.6%) |
| Monthly | 57 (15.0%) | 21 (36.8%) | 68 (10.9%) | 30 (44.1%) | 18 (10.4%) | 11 (61.1%) | 143 (12.1%) | 62 (43.4%) |
| Two Monthly | 49 (12.9%) | 11 (22.5%) | 85 (13.6%) | 39 (45.9%) | 21 (12.1%) | 9 (42.9%) | 155 (13.1%) | 59 (38.1%) |
| Three Monthly | 131 (34.5%) | 11 (8.5%) | 278 (44.3%) | 65 (23.4%) | 66 (38.2%) | 19 (28.8%) | 475 (40.3%) | 95 (20.0%) |
| Other | 44 (11.6%) | 21 (47.7%) | 68 (10.9%) | 37 (54.4%) | 14 (8.1%) | 11 (78.6%) | 126 (10.7%) | 69 (54.8%) |
| None (No treatment) | 70 (18.4%) | 6 (8.6%) | 96 (15.3%) | 10 (10.4%) | 45 (26.0%) | 4 (8.9%) | 211 (17.9%) | 20 (9.4%) |

## Discussion

Our study identified three distinct symptom-based subtypes within a predominantly PA confirmed population (76·4% with a formal PA diagnosis), including patients with B_12_ deficiency of uncertain etiology (21·8%). These subtypes, which we designate as High, Moderate, and Low Burden, were defined by cumulative symptom prevalence across multiple domains rather than diagnostic classification, addressing our objective to characterize clinical heterogeneity in this population. Each subtype demonstrated distinctive symptom profiles, demographic features, and treatment needs, supporting our hypothesis that symptom patterns, rather than diagnostic labels based on serological markers, better guide personalized management approaches (**Fig 2**). This represents the first systematic application of person-centered methodology in this patient population, extending beyond current literature which focuses primarily on cataloging symptoms rather than identifying subtypes.[15,17,25,32]

**Fig 2.**
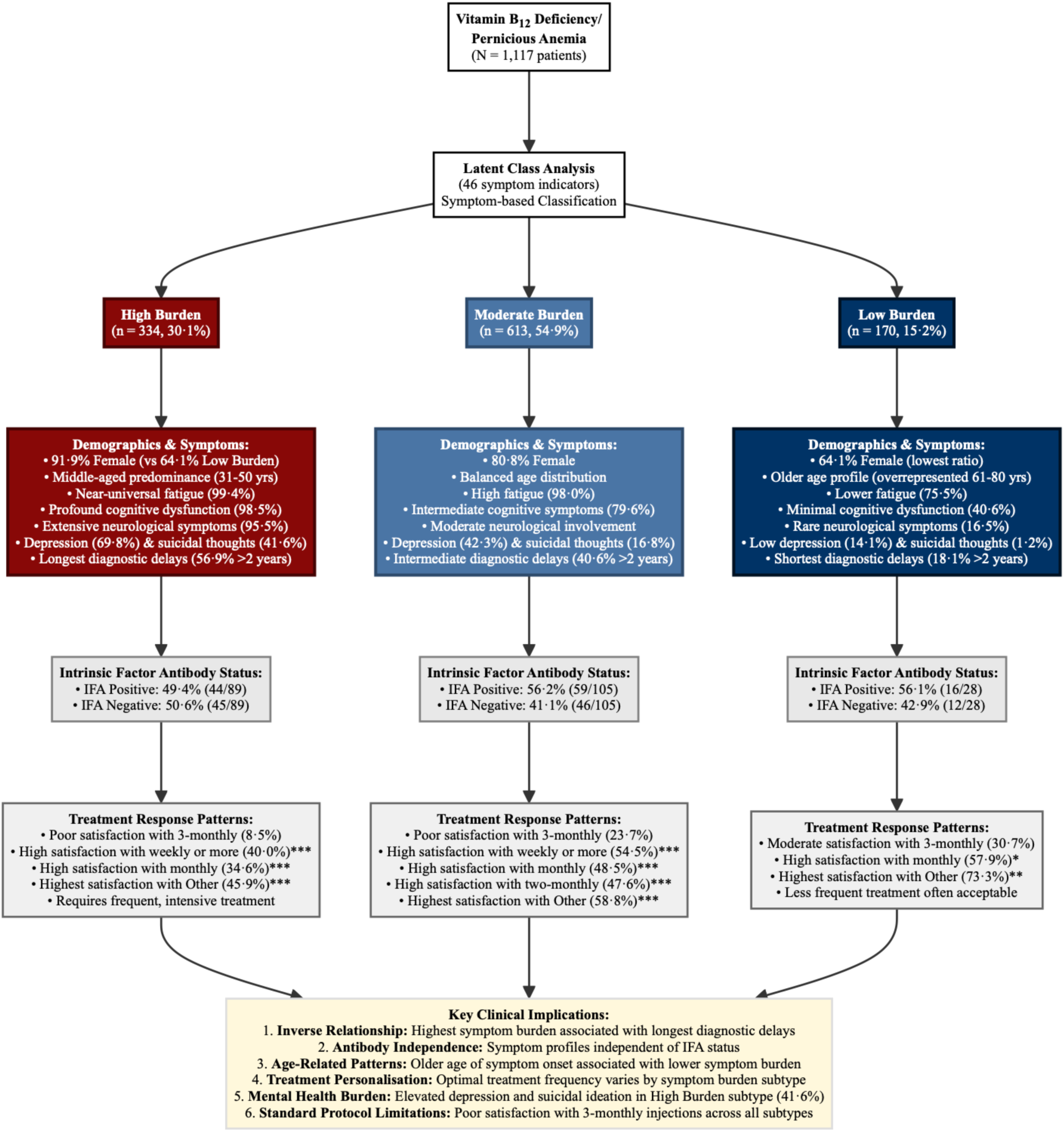
Latent Class Analysis of Vitamin B12 Deficiency/Pernicious Anemia Symptom Profiles and Clinical Characteristics. Study findings showing the three-subtype solution derived from latent class analysis of 46 indicators in 1,117 participants with vitamin B12 deficiency and/or pernicious anemia. The analysis identified three distinct symptom-based subtypes: High Burden, n=334 (29·9%), characterized by near-universal fatigue, profound cognitive dysfunction, and extensive neurological symptoms with the highest female predominance and longest diagnostic delays; Moderate Burden, n=613 (54·9%), showing intermediate symptom severity across all domains; and Low Burden, n=170 (15·2%), presenting with limited symptoms, older age profile, and shortest diagnostic delays. Intrinsic factor antibody (IFA) status did not significantly differentiate between subtypes (χ^2^=1·04, df=2, p=0·593; n=222 tested), while treatment response patterns varied by symptom burden subtype, with the High Burden subtype showing low satisfaction with three-monthly injection protocols and higher satisfaction with more frequent administration. Key clinical implications include the inverse relationship between symptom burden and diagnostic delay, the inadequacy of current standard treatment protocols across all subtypes, and the critical need for personalized, symptom-guided treatment approaches rather than antibody classification.

Previous literature describes both B_12_ deficiency and PA as clinically heterogenous, spanning neurological, hematological, psychiatric, and gastrointestinal manifestations.[7] Our analysis reveals that seemingly dissimilar symptoms cluster into distinct patterns giving three subtypes defined by cumulative symptom prevalence rather than individual symptom intensity. Whether these subtypes represent etiologically distinct phenotypes or severity strata along single disease continuum remains an open question for future studies. However, several patterns suggest these may be different disease processes rather than simply varying severity: the non-linear gradient of suicidal thoughts (41·6% in High Burden vs. 16·8% in Moderate Burden vs 1·2% in Low Burden), the older age profile of the Low Burden subtype, and differential multigenerational family history patterns are not straightforwardly explained by a simple severity spectrum. Prospective studies incorporating biomarkers, genetic profiling, and longitudinal symptom assessment will be necessary to determine whether these subtypes reflect different disease mechanisms or clinically meaningful points along a severity spectrum. This distinction has important implications for diagnostic classification and treatment stratification.

The High Burden subtype presented with overwhelming multisystem involvement, near-universal fatigue, waking tired, profound cognitive dysfunction including memory loss, poor concentration, and confusion, alongside prominent neurological manifestations with high rates of clumsiness, dizziness, and paresthesia. The Moderate Burden subtype, representing the largest group, demonstrated intermediate symptom burden; fatigue remained prevalent, but cognitive, neurological, and psychological symptoms were significantly less pronounced. The Low Burden subtype was an attenuated phenotype with a lower symptom burden across all domains. The shifting female-to-male ratio across subtypes (High Burden 13:1, Moderate Burden 5:1, Low Burden 3:1) also warrants follow-up, though these ratios may reflect PAS membership composition and volunteer bias rather than true population distributions.

The absence of association between IFA status and symptom burden (49·4%, 56·2%, 57·1%) must be interpreted alongside the well-documented technical limitations of IFA testing.[20] The approximately equal distribution of positive and negative results across all burden classes likely reflects the poor sensitivity of current antibody assays rather than meaningful biological distinctions. IFA testing suffers from substantial technical variability, with low sensitivity (as low as 27%) despite high specificity (>80%), making it unreliable for differentiating patient subgroups or predicting clinical outcomes.[20] This finding underscores that current serological markers are inadequate for stratifying patients or guiding treatment decisions. What remains clearer from our analysis is that symptom burden, independent of serological markers, strongly influences treatment needs and satisfaction, suggesting that clinical phenotyping may be a useful tool for development of more precise management plans.

While PA may take 10-12 years to progress from AIG to clinical presentation, it remains unknown whether diagnostic delays are associated with symptom burden.[8,40] Notably, we found an inverse relationship where individuals within the High Burden subtype, who are predominantly diagnosed before age 60, experienced significantly longer diagnostic delays (56·9% reporting more than 2 years before diagnosis) despite exhibiting widespread multisystem symptom profiles. This can be interpreted in several ways. First, lower symptom burden among older patients may reflect age-associated changes in gastric physiology, immune function, or nutrient absorption, possibly indicating slower disease progression and milder symptomatology burden; and diagnostic attribution bias, whereby symptoms in older patients are more readily ascribed to normal aging rather than investigated as potential B_12_ deficiency, potentially compounding delay recognition. Second, High Burden B_12_ deficiency in younger patients might represent a distinct, more aggressive phenotype, driven by stronger autoimmune and/or genetic factors.[41,42] The higher rates of multigenerational family history in the High Burden subtype support this interpretation, suggesting stronger genetic predisposition potentially linked to HLA-DRB103, HLA-DRB104, and other autoimmune susceptibility alleles.[41] Beyond these interpretations, B_12_-dependent enzymatic pathways offer a plausible biological basis for symptom heterogeneity observed across subtypes. Deficiency in adenosyl-B_12_ leads to MMA accumulation, while methyl-B_12_ deficiency results in homocysteine elevation and impaired methylation affecting myelin synthesis and neurotransmitter production.[18,43] Whether differential disruption of these pathways underlies the symptom burden gradient remains to be established, though the neurological and neuropsychiatric manifestations of the High Burden subtype are consistent with this possibility.[44] This subtype also had the highest rate of self-reported folic acid deficiency (32·2%), which, while not clinically verified in this study, raises the possibility that micronutrient deficiencies beyond B_12_ may contribute to symptom burden in this group.[45,46] However, folate supplementation in inadequately treated B_12_ deficiency may mask hematological signs of deficiency while accelerating neurological deterioration.[47] Given the pronounced neurological burden already observed in this subtype, ensuring adequate B_12_ replacement before or alongside folate supplementation is essential.[48,49]

Across all three symptom subtypes, we identified significant associations between treatment frequency and satisfaction. The highest satisfaction rates were observed with alternative treatment approaches outside standard schedules (Other: 45·9%-73·3%), followed by weekly or more frequent injections (40·0%-62·5%), then monthly treatments (34·6%-57·9%). Standard three-month injections showed consistently poor satisfaction across all subtypes (8·5% - 30·7% satisfaction), supporting the need for more symptom-tailored treatment protocols. Kornic et al. reported that more frequent injection intervals were associated with greater patient-reported symptom improvement.[50] Our demonstration that optimal treatment frequency varies by symptom-burden subtype extends prior research by showing that symptom phenotypes drive differential treatment response patterns and provides supportive evidence for exploring personalized, symptom-based therapeutic approaches. It should be noted, however, that neurological and psychiatric symptoms are known to show greater resistance to B_12_ supplementation than hematological manifestations, meaning that lower satisfaction in the High Burden subtype may partly reflect treatment-resistant symptomology rather than inadequate administration frequency alone.[51] This limits causal interpretation of the satisfaction gradient and underscores the need for pragmatic trials examining biological response across symptom subtypes

The mental health gradient across subtypes warrants particular attention. The High Burden subtype showed remarkably high rates of depression (69·8%) and suicidal ideation (41·6%), substantially exceeding both Moderate Burden (42·3%; 16·8%) and Low Burden (14·1%; 1·2%) subtypes. These observations likely reflect at least two complementary mechanisms. First, B_12_ deficiency creates direct biological vulnerability to psychiatric manifestations through impaired neurotransmitter synthesis and disrupted methylation pathways, effects that may be most pronounced in those with the greatest overall metabolic burden.[47,52,53] Second, prolonged diagnostic delay imposed substantial situational and existential load on top of this biological vulnerability, encompassing feelings of medical invalidation, grief over lost functional years, and compounding social stressors such as occupational disruption and financial strain.[54,55] These mechanisms are not mutually exclusive, biological vulnerability and situational burden are likely to interact and amplify one another, particularly among those experiencing the longest delays. The predominantly female composition of the High Burden subtype (91·4%) adds a further dimension. Evidence documents that women are disproportionately subject to medical gaslighting, the systemic attribution of physical symptoms to psychological cause, which may deepen both diagnostic delay and psychological distress through cumulative invalidating clinical encounters.[56] This pattern likely compounds the biological vulnerability and situational burden already experienced by this subtype, further contributing to the mental health gradient observed across subtypes. This suggests that High Burden B_12_ deficiency warrants integrated clinical attention addressing both underlying deficiency and its psychological sequelae, particularly given the high reported rates of suicidal ideation.[57] Further research at the intersection of B_12_ deficiency and mental health, including neurobiological mechanisms and adjunctive psychiatric treatment approaches, is warranted.

Addressing the treatment frequency gradient and the mental health burden identified across subtypes requires systematic changes to clinical practice. This aligns with principles of the CCM, a framework stating that effective management of chronic conditions requires prepared, proactive healthcare teams working collaboratively with informed, engaged patients rather than reactive, crisis-driven care.[31] The 2024 NICE guideline, recommends titrating intramuscular B_12_ to symptom severity or response rather than following standardized protocols, representing a significant policy shift toward this patient-centered model. However, the guidelines provide limited guidance on how clinicians should operationalize symptom titration in practice. Our identification of distinct symptom subtypes, with differential treatment needs, provides support for an evidence-based framework for operationalizing the NICE recommendations through CCM-informed approaches. For example, clinical information systems could track symptom profiles and treatment responses over time, delivery system redesign could enable flexible dosing protocols tailored to symptom subtype, and self-management support could help patients monitor their symptoms, self-adjust their injection frequency, and communicate changes to providers.[30,58] The CCM framework is particularly well-suited to this population given evidence that integrated management of depression alongside the underlying chronic condition produces better outcomes than treating each in isolation, an approach directly relevant to the High Burden subtype where biological and psychosocial contributors to depression likely to coexist.

This study has notable strengths. We used data-driven agnostic methods to identify latent subgroups and examined how symptom profiles predict treatment response, moving beyond the descriptive satisfaction rates or treatment preferences reported in previous research.[24,26,50] By including patients across a diagnostic spectrum rather than restricting analysis to confirmed PA cases, we examined symptom patterns independent of diagnostic labels. Importantly, participants were asked to report symptoms experienced prior to diagnosis rather than current symptoms, anchoring the analysis to the pre-diagnostic presentation most relevant for informing clinical recognition and treatment stratification. Our comprehensive assessment of symptoms across multiple domains provides a holistic view of disease burden not captured by traditional clinical parameters. Additionally, the consistency of findings across different diagnostic categories and demographic subgroups enhances generalizability within similar patient advocacy populations.

Several limitations merit consideration. Our sample consisted exclusively of PAS members, who represent a highly engaged patient population that may differ systematically from broader B_12_-deficient populations, potentially overrepresenting those with High Burden symptoms or treatment dissatisfaction and limiting applicability to community-based clinical settings.[24,26,50] The predominance of female participants (81·6%), and the limited ethnic diversity within PAS membership prevents examination of potential gender and ethnic disparities in PA prevalence or symptom severity, an important area for future investigation in more diverse populations.

The survey was administered between 2010 and 2012, prior to the 2024 revision of the NICE guidelines on B_12_ deficiency and PA; the diagnostic and treatment landscape available to clinicians at the time differed substantially from current practice, and participants’ experiences should be interpreted in this historical context. Data were self-reported rather than extracted from medical records, which introduces the possibility of recall bias and prevents independent verification of diagnoses or treatment details from clinics. The survey lacked formal psychometric validation, and our binary treatment satisfaction measure may not capture nuanced responses. Finally, the cross-sectional design prevents examining symptom evolution or establishing causal relationships, limiting our ability to determine whether symptom subtypes represent stable phenotypes or disease progression stages, or might in part reflect conditions other than B_12_ deficiency.

In conclusion, our identification of three distinct symptom-based subtypes in B_12_ deficiency and PA represents a shift from diagnostic category-driven to symptom pattern-guided approaches. These subtypes, characterized by different symptom burdens, demographic profiles, and treatment response patterns, provide more clinically relevant information than a differential diagnosis of either B_12_ deficiency or PA. The striking finding that standard three-monthly injection protocols showed poor satisfaction across all subtypes (8·5% to 30·7%), while frequency requirements varied systematically by symptom burden subtype, has immediate implications for clinical practice. These findings support recent NICE guidelines advocating symptom-guided treatment frequency and suggest that many patients currently receive suboptimal care under standardized protocols. For clinicians, these results argue for implementation of systematic symptom assessment, personalized treatment schedules based on symptom subtype along with heightened vigilance for mental health complications, particularly in severely affected patients. At the policy level, this evidence supports replacing rigid treatment protocols with flexible, patient-centered approaches that recognize symptom heterogeneity and by empowering patients as active partners in care decisions, potentially improving outcomes while reducing healthcare utilization from symptom relapses and crisis interventions.

## Data Availability

The datasets generated and/or analyzed during the current study are not publicly available as the consent provided by participants does not include permission for public availability.

## Abbreviations

α: Alpha; significance threshold
aBIC: Sample-size adjusted Bayesian Information Criterion
AIC: Akaike Information Criterion
AIG: Autoimmune gastritis
B_12_: Vitamin B12 (cobalamin)
BIC: Bayesian Information Criterion
CCM: Chronic care model
COVID-19: Coronavirus disease 2019
df: egrees of freedom
DNA: Deoxyribonucleic acid
EM: Expectation-maximization
FIML: Full information maximum likelihood
HLA: Human leukocyte antigen
IFA: Intrinsic factor antibodies
LCA: Latent class analysis
LMR: LRT Lo-Mendell-Rubin adjusted likelihood ratio test
MLR: Maximum likelihood ratio
MMA: Methylmalonic acid
NHS: National Health Service
NICE: National Institute for Health and Care Excellence
PA: Pernicious anaemia/anemia
PAS: Pernicious Anaemia Society
PCA: Parietal cell antibodies
QoL: Quality of life
STROBE: Strengthening the Reporting of Observational Studies in Epidemiology
UK: United Kingdom
χ²: Chi-square
ρ: Spearman rank correlation coefficient

## Supplementary Methods

### Survey Development

The survey instrument was originally developed by the Pernicious Anaemia Society (PAS) in collaboration with a general practitioner (GP) using SurveyMonkey software between August 2010 and November 2012. The survey was designed based on clinical expertise and extensive patient input through the PAS membership, following identification of widespread patient concerns regarding diagnostic delays and treatment dissatisfaction within the patient community.

The comprehensive survey instrument consisted of 38 questions organized into four main sections (**S1 Table**): 1. Demographics (Part A) is 11 questions covering basic demographic information including sex, age at diagnosis, ethnicity, eye and hair color, family history of pernicious anemia, and dietary patterns (vegetarian/vegan status). 2. Symptom Assessment (Part B) covers systematic documentation of symptoms experienced prior to diagnosis across seven physiological domains: general symptoms (10 items), neurological symptoms (13 items), respiratory symptoms (1 item), cardiovascular symptoms (1 item), gastrointestinal symptoms (6 items), genitourinary symptoms (1 item), and emotional symptoms (4 items). 3. Comorbid conditions (Part C) comprised of 21 questions identifying coexisting medical conditions potentially associated with pernicious anemia across eight clinical areas: general, neurological, dermatologic, cardiovascular, musculoskeletal, endocrine, gastrointestinal, and emotional conditions. 4. Clinical assessment and treatment (Part D) are10 questions assessing diagnostic testing procedures, treatment modalities (type, frequency, and delivery method), treatment satisfaction, and patient experience with healthcare providers

Question formats included binary yes/no responses, single-selection multiple choice, multi-selection options (e.g., “tick all that apply”), and ordinal scales. For treatment satisfaction assessment, participants rated their medical care using a 10-point ordinal scale ranging from “Excellent” to “Unreasonable.”

### Statistical Software and Technical Specifications

All latent class analyses were conducted using Mplus Version 8·0 with full information maximum likelihood estimation using the robust maximum likelihood ratio (MLR) estimator to handle non-normality in binary indicators.[1] R Statistical Software (version 4·0·3; R Core Team 2021) was used for descriptive statistics, post-hoc analyses, and data visualization.[2]

### Missing Data Pattern Analysis

The percentage of missing values was calculated for all variables included in the survey. All LCA candidate variables demonstrate complete or near-complete data across the analytic sample (N=1,117). Missing data were only observed in post-hoc outcome variables (age when symptoms first started, duration of symptoms before diagnosis, and treatment satisfaction). Participants with missing post-hoc data were retained in the LCA but excluded listwise from the relevant post-hock analysis.

### Recall Bias Assessment

To address potential recall bias in retrospective symptom reporting, Spearman rank correlations were calculated between symptom duration before diagnosis and individual symptom endorsement. Symptom duration was measured categorically (1 week to >10 years) and converted to months for analysis. While statistical significance was assessed at α=0·05, correlations were considered practically significant only if |ρ| ≥ 0·30, following established guidelines for clinical research.

### Latent Class Analysis Model Specifications

Latent class models were estimated using the expectation-maximization (EM) algorithm with 1,000 random starting values to ensure identification of the global maximum likelihood solution. Models were considered converged when the change in log-likelihood between iterations was less than 1×10⁻⁶. To address potential local maxima, we examined the 100 best log-likelihood values from the random starts and verified that the final solution was replicated across multiple starting values.

The selection of indicators for latent class analysis followed established methodological guidelines to ensure model stability and interpretability. From the original survey instrument, systematic selection criteria were applied including a prevalence threshold where variables were included only if endorsed by ≥10·0% of the analytic sample (n≥112), following recommendations by Sinha et al. (2021) to prevent identification of classes based solely on rare indicators.[3]

Variables were retained if they represented established manifestations of B_12_ deficiency or PA based on clinical literature review. This approach resulted in 46 indicators being included in the analysis, comprising three demographic indicators, 35 symptoms indicators across multiple physiological domains, and eight comorbid condition indicators.

### Model Selection Strategy

Model selection was based on multiple fit indices examined simultaneously, including the Akaike Information Criterion (AIC), Bayesian Information Criterion (BIC), and sample-size adjusted BIC (aBIC), where lower values indicate better fit, with BIC providing stronger penalty for model complexity. Statistical tests included the Lo-Mendell-Rubin likelihood ratio test (LMR-LRT), which tests whether a k-class model fits significantly better than a (k-1)-class model, and entropy, which measures classification quality with values closer to 1·0 indicating clearer class separation.

Models with 2–10 classes were estimated and compared through visual examination of information criteria plots to identify points of diminishing returns in model fit improvement.[4] The final model was selected based on the convergence of fit indices, LMR-LRT significance patterns, adequate and stable class sizes, and theoretical interpretability of the resulting subtypes.

## Post-Hoc Analyses

### Age Distribution Analysis

Age when symptoms first started was analyzed as a categorical variable in nine ranges (0-10, 11-20, 21-30, 31-40, 41-50, 51-60, 61-70, 71-80, 81-90) and evaluated using chi-square analysis to test for associations with symptom burden type. To identify which specific age and subtype combination drove any observed associations, standardized residuals were calculated for each age group and subtype combinations; absolute values above 1·96, corresponding to p<0·05 under the standard normal distribution, were treated as statistically significant and are presented in **S4 Fig**. The full distribution of participants across age groups and subtypes is presented in **S4 Table**. Participants with missing data for age when symptoms first started were excluded listwise from the analysis (n=1,013, 90·7% of the analytic sample).

### Symptom Duration Analysis

Duration of symptoms before diagnosis was collected using 11 pre-specified ordered response categories (1 week, 2 weeks, 1 month, 2 months, 3 months, 6 months, 1 year, 2 years, 5 years, 10 years, and more than 10 years). For visualization and analysis of diagnostic delays, duration categories were collapsed into broader clinically relevant groups: less than 6 months, 6 months to 2 years, and more than 2 years (**S5 Fig**). The full 11-category distribution is presented in **S6 Fig**. Chi-square analysis with Cramér’s V effect size was used to examine associations between symptom duration and symptom burden subtype for both the collapsed (df=4) and full (df=20) category analyses. Participants with missing data for symptom duration were excluded listwise from both analyses (71 of 1,117 participants; n=1,046, 93·6% of the analytic sample).

### Treatment Frequency and Satisfaction Analysis

Treatment frequency and satisfaction rates were examined across symptom burden subtypes among participants with a recorded treatment frequency and satisfaction response (n=927). Frequency categories were collapsed where appropriate: weekly or more (daily, more than once a day, and weekly), monthly, two-monthly, three monthly and other. Participants with missing treatment frequency data (n=155, 13·9% of the analytic sample) or missing satisfaction responses (n=37) were excluded listwise from all treatment analyses. Fisher’s exact tests with 10,000 Monte Carlo simulations were used to compare satisfaction rates for each category against the three-monthly injections as the reference category, reflecting the standard NHS treatment schedule. This approach was selected over chi-squared testing as some cells had small expected counts.

Free-text responses in the “Other” category (n=125) were examined separately. Each response was coded by injection interval in weeks and categorized into five groups: loading phase (initial loading doses without an established maintenance frequency), very frequent (every two weeks or less, including daily and alternate day), extended standard (every 3-11 weeks), extended interval (every 12 weeks or more), and no treatment or pre-treatment (participants not yet receiving or unable to receive maintenance injections). Satisfaction rates were calculated within each frequency type and subtype among participants providing a Yes or No satisfaction response.

The “Weekly or more” category in Table 3 and the “Very frequent (:≤ 2 weeks)” category in **S5 Table** are mutually exclusive groups. Table 3 presents participants who selected structured frequency options from the survey response list (Daily, More than once a day, or Weekly), pooled into a single “Weekly or more” category due to small cell sizes. **S5 Table** presents only participants who selected “Other” and provided a free-text description of their injection frequency; these participants did not select any structured frequency option. The “Very frequent’ category in Supplementary Table 5 therefore reflects participants whose self-described frequency was equivalent to weekly or more frequent administration, but who chose to describe it in free text rather than selecting a structured response option. The two groups are non-overlapping.

### Statistical Significance and Multiple Comparisons

Statistical significance was set at α = 0·05 for all analyses. P-values are reported to two significant figures (capped at four decimal places) or as p<0·0001 for highly significant results. Given the exploratory nature of latent class analysis and the large number of post-hoc comparisons, emphasis was placed on effect sizes and clinical interpretability rather than strict multiple comparison corrections.

## Supplementary Results

### Missing Data Analysis

Complete data were available for all 46 LCA indicator variables across the full analytic sample (N=1,117). Missing data were only observed in post-hoc outcome variables: age when symptoms frost started (9·3%, n=104), duration of symptoms before diagnosis (6·4%, n=71), and treatment satisfaction (8·1%, n=90). Participants with missing post-hoc data were excluded listwise from the relevant post-hoc analyses (**S1 Fig**).

Missing data analysis revealed minimal missingness across variables included in the latent class analysis. The majority of key variables showed <1·0% missing data, supporting the robustness of the classification, while variables exceeding 10% missingness were excluded from the LCA indicator set. The optional question regarding children having pernicious anemia showed 97·9% missingness, reflecting the survey design where this question was not applicable to many participants. Chi-square tests confirmed that missing data patterns were not significantly associated with participant demographics (p>0·05), supporting the assumption that data were missing at random within the context of observed variables (**S1 Fig**).

### Recall Bias Assessment

Spearman rank correlations between symptom duration and individual symptom endorsement were examined to assess potential recall bias in retrospective symptom reporting (**S2 Table, S2 Fig**). While 33 of 35 symptoms (94·3%) showed statistically significant correlations with symptom duration (p<0·05), the magnitude of these correlations was uniformly weak, ranging from -0·032 to 0·205. No correlations exceeded the threshold for practical significance (|ρ|≥0·30), suggesting that recall bias had minimal impact on symptom reporting and supporting the validity of retrospective symptom data for latent class analysis. The highest correlations were observed for brittle nails with ridges (ρ=0·205), shoulder bumps (ρ=0·195), and vertigo (ρ=0·193), but even these remained well below thresholds for clinical concern.

### Latent Class Analysis Model Selection and Diagnostics

Latent class models with 2-10 classes were systematically evaluated using multiple information criteria (**S3 Table, S3 Fig**). The 3-class solution emerged as optimal based on several converging indicators. The 3-class model showed substantial improvement over the 2-class solution across all information criteria, with AIC decreasing from 56733 to 55866, BIC from 57200 to 56568, and aBIC from 56904 to 56123. While the 4-class model achieved a slightly lower BIC (56506), the improvement was minimal and not statistically significant. The LMR-LRT confirmed significant improvement from 2 to 3 classes (p<0·0001) but showed no significant improvement from 3 to 4 classes (p=0·51), supporting the 3-class solution.

The 3-class model demonstrated high entropy (0·86), indicating clear separation between classes and robust participant classification. The solution produced three distinct and interpretable symptom burden subtypes with adequate sample sizes (High Burden; n=334, Moderate Burden; n=613, Low Burden; n=170. The final 3-class solution demonstrated robust statistical properties, with model convergence achieved across all 1,000 random starting values and consistent replication of the highest log-likelihood value, confirming identification of the global maximum likelihood solution.

### Age Distribution Analysis

Significant differences in age when symptoms first started were observed across the three symptom burden subtypes (χ²=37·07, df=16, p=0·002, Cramér’s V=0·14; **S4 Fig**, **S4 Table**). The Low Burden subtype showed overrepresentation in older age groups, particularly ages 71-80 (7·7% vs. 3·2% overall) and 61-70 (13·5% vs. 8·4% overall). The High Burden subtype showed overrepresentation in the 31-40 age group (28·5% vs 24·2% overall) and underrepresentation in the 71-80 age group (1·0% vs. 3·2% overall). The Moderate Burden subtype demonstrated the most balanced age distribution, closely matching the overall sample demographics across all age categories.

### Symptom Duration and Diagnostic Delay Analysis

Analysis of symptom duration before diagnosis revealed an inverse association between symptom burden and diagnostic timeliness (**S5 Fig, S6 Fig**), where participants with the highest symptom burden also experienced the longest pre-diagnosis periods. The High Burden subtype showed particularly prolonged pre-diagnosis periods, with 56·9% of participants experiencing symptoms for more than two years before diagnosis. Conversely, the Low Burden subtype was diagnosed more rapidly, with 21·8% receiving a diagnosis within six months of symptom onset. A clear gradient was observed across subtypes, with diagnostic delays systematically increasing with symptom burden (χ²=115·47, df=4, p<0·001, Cramér’s V=0·23, n=1,046). The full 11-category distribution confirmed the pattern (χ²=149·59, df=20, p<0·001, Cramér’s V=0·27), though interpretation should be made with caution as some cells had small expected counts.

### Treatment Frequency and Satisfaction Analysis

Treatment frequency and satisfaction data were available for 927 of 1,117 participants (82·8%; 155 excluded for missing frequency data, 37 for missing satisfaction response). Three-monthly injections were the most common treatment frequency across all subtypes (High Burden: 42·8%; Moderate Burden: 51·1%; Low Burden: 54·3%),yet showed consistently low satisfaction rates (High Burden: 8·5%; Moderate Burden: 23·7%; Low Burden: 31·1%).

In the High Burden subtype, weekly or more (40·0%, p<0·001), monthly (34·6%, p<0·001), and Other (45·9%, p<0·001) frequencies were all associated with significantly higher satisfaction than three-monthly injections. Two-monthly injections approached but did not reach significance (20·5%, p=0·063). In the Moderate Burden subtype, all frequencies showed significantly higher satisfaction than three-monthly injections (weekly or more: 54·5%, p=0·001; monthly: 48·5%, p<0·001; two-monthly: 47·6%, p<0·001; Other: 58·8%, p<0·001). The Low Burden subtype showed a different pattern, with only monthly (57·9%, p=0·037) and Other (73·3%, p=0·002) frequencies reaching significance compared to three-monthly injections; weekly or more (62·5%, p=0·115) and two-monthly (47·6%, p=0·197) did not differ significantly from three-monthly injections in this subtype.

Analysis of the 125 participants selecting “Other” treatment frequency revealed distinct patterns acroass subtypes (Supplemental Table S5). Very frequent injections (every two weeks or less) were associated with the highest satisfaction rates in the High Burden (77·8%, n=9) and Moderate Burden (81·8%, n=12) subtypes. No High Burden participants reported extended interval injections (every 12 weeks or more). Among the five High Burden with no established treatment, satisfaction was 0·0%, reflecting unmet treatment need in the highest symptom burden group. Loading phase participants shows moderate satisfaction across High Burden (42·9%) and Moderate Burden (42·9%) subtypes, consistence with treatment being in an early transitional stage.

The overlap in apparent frequency between the “Weekly or more” category in Table 3 and the “Very frequent (≤ 2 weeks)” category in Supplementary Table 5 reflects a survey design feature rather than double-counting. Participants in Table 3 selected structured frequency options, while those in Supplementary Table 5 selected “Other” and described equivalent frequencies in free text. Combined, these groups suggest that high-frequency administration, whether formally categorized or self-described, is associated with higher satisfaction rates, particularly in the High Burden subtype.

## Supplementary Tables

**S1 Table:**
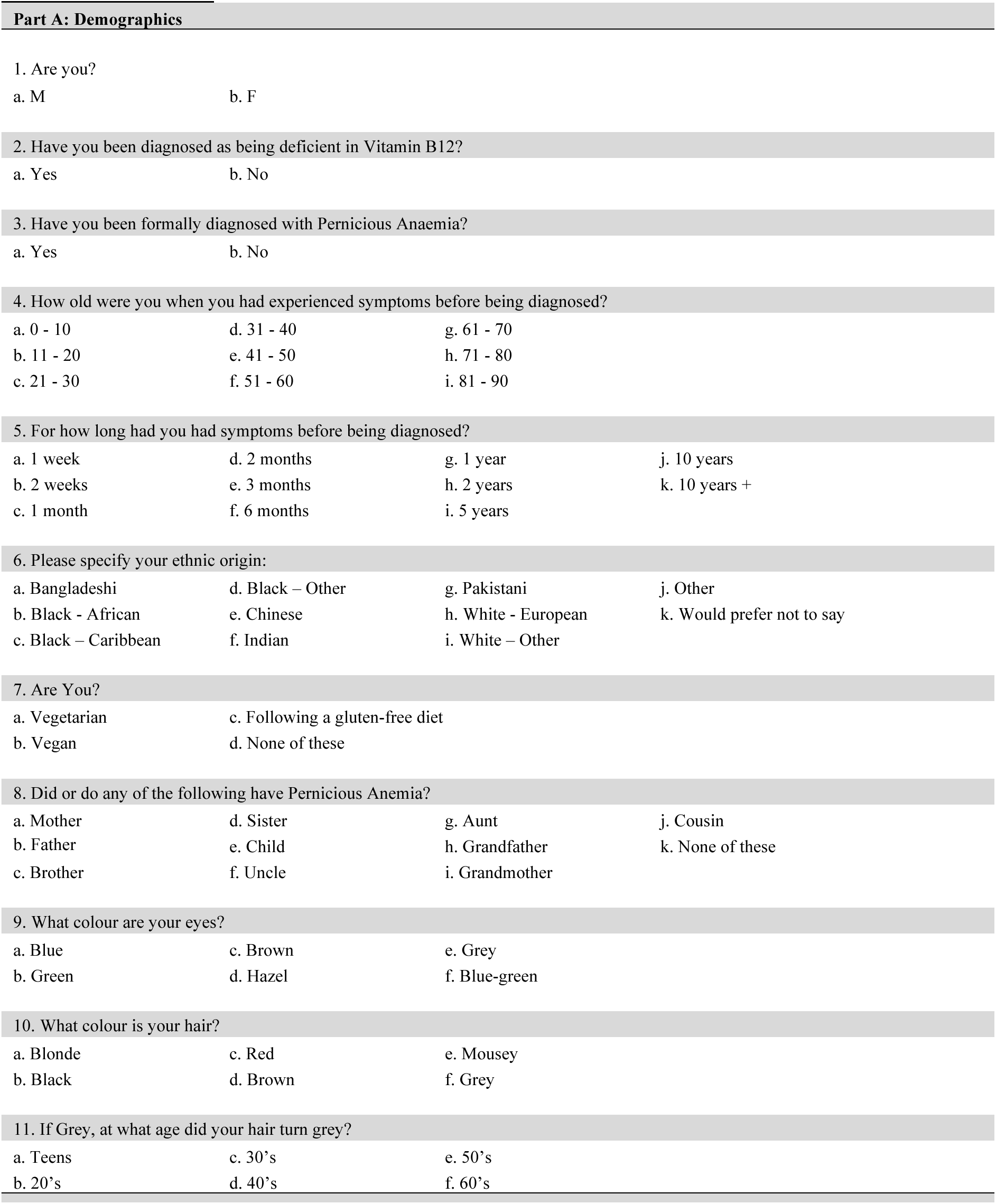
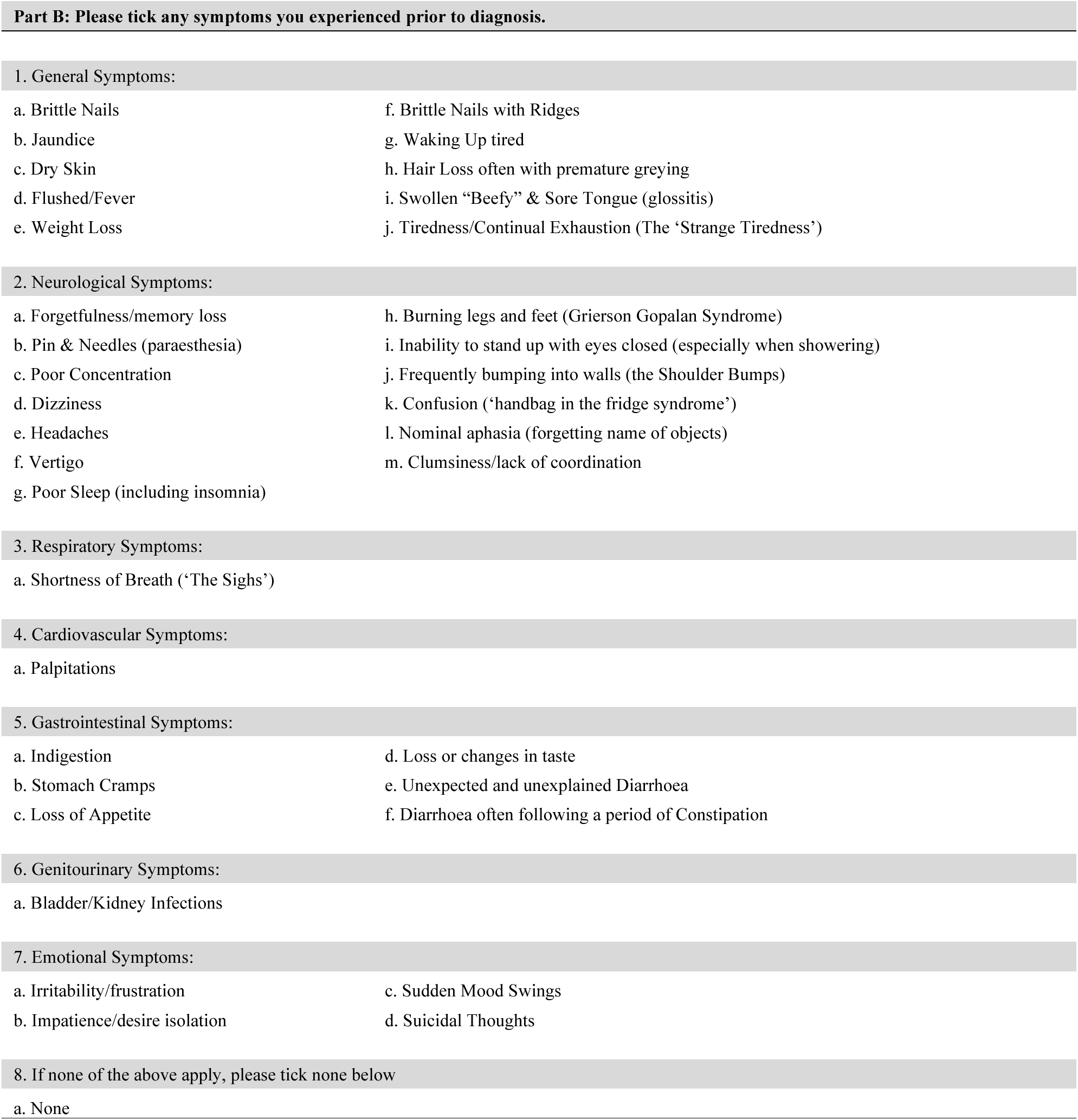
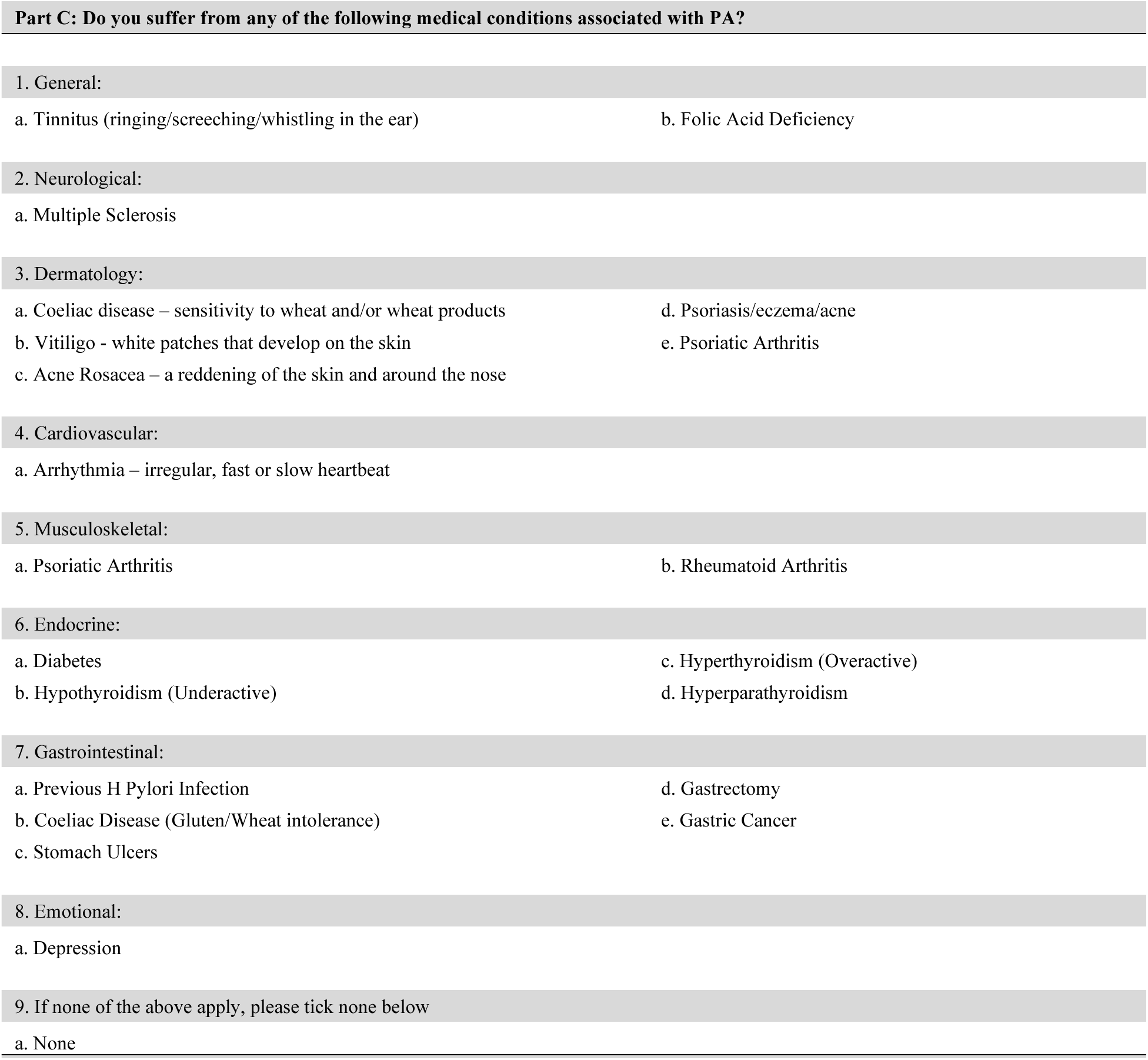
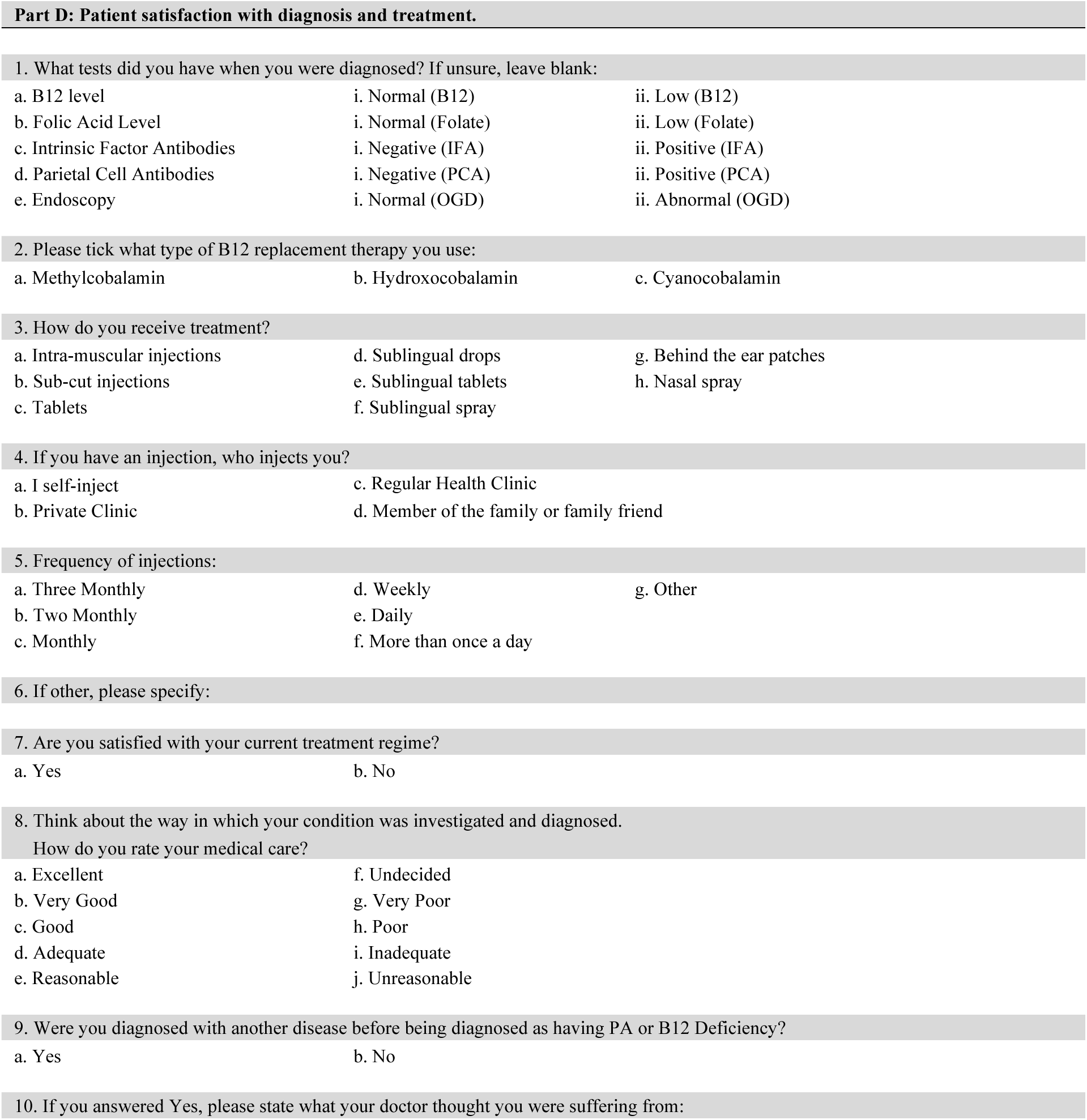
Pernicious Anaemia Society survey instrument (August 2010 - November 2012). The survey comprises four sections: Part A collects demographic information; Part B documents symptoms experienced prior to diagnosis; Part C identifies comorbid conditions potentially associated with pernicious anaemia; and Part D assesses diagnostic testing, treatment modalities, and patient satisfaction with care.

**S2 Table:** Spearman correlation coefficients between duration of symptoms before diagnosis and individual symptom endorsement (N=1,117). Values represent Spearman correlation coefficients (ρ) and corresponding p-values for 35 symptoms. Duration of symptoms before diagnosis was measured categorically (1 week to >10 years) and converted to months for analysis. Statistically significant correlations (p<0·05) were observed for 33 of 35 symptoms; however, all coefficients were weak in magnitude (range -0·032 to 0·205). suggesting minimal practical impact of recall bias on symptom reporting.

| Symptom | $\rho$ | p-value |
| --- | --- | --- |
| Tiredness | 0.076 | 0.014 |
| Hair loss or greying | 0.157 | <0.001 |
| Brittle nails | 0.092 | 0.003 |
| Brittle nails with ridges | 0.205 | <0.001 |
| Weight loss | -0.032 | 0.302 |
| Wake up tired | 0.125 | <0.001 |
| Dry skin | 0.138 | <0.001 |
| Flushes fever | 0.124 | <0.001 |
| Glossitis | 0.169 | <0.001 |
| Poor concentration | 0.115 | <0.001 |
| Pins & needles (paresthesia) | 0.126 | <0.001 |
| Confusion | 0.156 | <0.001 |
| Nominal aphasia | 0.179 | <0.001 |
| Clumsiness | 0.156 | <0.001 |
| Headaches | 0.163 | <0.001 |
| Grierson Gopalan Syndrome (burning legs and feet) | 0.159 | <0.001 |
| Forgetfulness/memory loss | 0.126 | <0.001 |
| Poor sleep | 0.103 | 0.001 |
| Dizziness | 0.160 | <0.001 |
| Shoulder bumps | 0.195 | <0.001 |
| Can't stand eyes closed | 0.144 | <0.001 |
| Vertigo | 0.193 | <0.001 |
| Shortness of breath (The sighs) | 0.113 | <0.001 |
| Palpitations | 0.184 | <0.001 |
| Indigestion | 0.181 | <0.001 |
| Loss of appetite | 0.043 | 0.168 |
| Taste loss or change | 0.069 | 0.026 |
| Stomach cramps | 0.148 | <0.001 |
| Diarrhea | 0.169 | <0.001 |
| Diarrhea alternating with constipation | 0.154 | <0.001 |
| Bladder/kidney infections | 0.104 | 0.001 |
| Irritability | 0.159 | <0.001 |
| Impatience | 0.163 | <0.001 |
| Sudden mood swings | 0.086 | 0.005 |
| Suicidal thoughts | 0.176 | <0.001 |
| **S2 Table: Spearman correlation coefficients between duration of symptoms before diagnosis and individual symptom endorsement (N=1,117).** Values represent Spearman correlation coefficients ($\rho$) and corresponding p-values for 35 symptoms. Duration of symptoms before diagnosis was measured categorically (1 week to >10 years) and converted to months for analysis. Statistically significant correlations ($p<0.05$) were observed for 33 of 35 symptoms; however, all coefficients were weak in magnitude (range -0.032 to 0.205), suggesting minimal practical impact of recall bias on symptom reporting. | | |

**S3 Table:** Latent Class Analysis model fit statistics for 2-10 class solutions (N=1,117). BIC, AIC< and aBIC are presented alongside entropy values and Lo-Mendell-Rubin likelihood ratio test (LMR-LRT) p-values for each solution. While AIC and aBIC continue to decrease across all models, BIC shows a clear inflection points at the 3-class solution, supporting selection of this model as the optimal balance between fit and parsimony. Entropy remained high across all solutions (range 0·80-0·87), indicating good classification quality throughout. The LMR-LRT indicated significant improvement adding a third class (p < 0·0001) but non-significant improvement for the 4-class (p = 0·51), further supporting a 3-class solution. Class sizes for the selected 3-class solution were adequate (High Burden: n=334, Moderate Burden: n=613, Low Burden: n=170). BIC = Bayesian Information Criterion; AIC = Akaike Information Criterion; aBIC = sample-size adjusted BIC; LMR-LRT = Low-Mendell-Rubin adjusted Likelihood ratio test; ·· = not applicable.

| Model | BIC | AIC | aBIC | Entropy | LMR | Class Size |  |  |  |  |  |  |  |  |  |
| --- | --- | --- | --- | --- | --- | --- | --- | --- | --- | --- | --- | --- | --- | --- | --- |
| 2 | 57200·10 | 56733·39 | 56904·71 | 0·87 | <0·0001 | 520 | 597 | · | · | · | · | · | · | · | · |
| 3 | **56568·60** | **55866·02** | **56123·92** | **0·86** | **<0·0001** | **170** | **334** | **613** | · | · | · | · | · | · | · |
| 4 | 56506·08 | 55567·64 | 55912·12 | 0·84 | 0·51 | 125 | 397 | 474 | 121 | · | · | · | · | · | · |
| 5 | 56538·74 | 55364·43 | 55795·49 | 0·80 | 0·29 | 183 | 129 | 343 | 144 | 318 | · | · | · | · | · |
| 6 | 56624·53 | 55214·48 | 55732·00 | 0·80 | 0·45 | 185 | 139 | 130 | 139 | 302 | 222 | · | · | · | · |
| 7 | 56756·29 | 55110·26 | 55714·48 | 0·82 | 0·40 | 149 | 125 | 109 | 302 | 191 | 106 | 135 | · | · | · |
| 8 | 56893·95 | 55012·05 | 55702·85 | 0·82 | 0·76 | 101 | 248 | 123 | 111 | 92 | 142 | 165 | 135 | · | · |
| 9 | 57039·38 | 54921·61 | 55698·99 | 0·82 | 0·81 | 118 | 100 | 126 | 229 | 98 | 131 | 150 | 137 | 28 | · |
| 10 | 57191·91 | 54838·28 | 55702·24 | 0·83 | 0·80 | 111 | 120 | 159 | 28 | 107 | 135 | 97 | 118 | 149 | 93 |

**S4 Table:** Age when symptoms first started across symptom burden subtypes (n=1013). Values represent number of participants (percentage within subtype) for each 10-year age category. Participants with missing data for age when symptoms first started were excluded listwise (104 of 1,117 participants). Significant differences in age distribution were observed across classes (χ2 = 37・07, df = 16, p=0・002, Cramer’s V=0・14). The Low Burden subtype showed significant overrepresentation in older age groups, particularly ages 61-70 (13・5% vs. 8・4% overall) and 71-80 (7・7% vs. 3・2% overall). The High Burden subtype showed overrepresentation in the 31-40 age group (28・5% vs 24・2% overall) and underrepresented in the 71-80 age group (1・0% vs. 3・2% overall). The Moderate Burden subtype demonstrated the most balanced age distribution across all age categories

| Age Group | High Burden | Moderate Burden | Low Burden |
| --- | --- | --- | --- |
| 0-10 | 1 (0.3%) | 3 (0.5%) | 2 (1.3%) |
| 11-20 | 7 (2.4%) | 15 (2.7%) | 1 (0.6%) |
| 21-30 | 45 (15.3%) | 82 (14.6%) | 19 (12.3%) |
| 31-40 | 84 (28.5%) | 128 (22.7%) | 33 (21.3%) |
| 41-50 | 91 (30.8%) | 160 (28.4%) | 36 (23.2%) |
| 51-60 | 46 (15.6%) | 104 (18.5%) | 27 (17.4%) |
| 61-70 | 17 (5.8%) | 47 (8.3%) | 21 (13.5%) |
| 71-80 | 3 (1.0%) | 17 (3.0%) | 12 (7.7%) |
| 81-90 | 1 (0.3%) | 7 (1.2%) | 4 (2.6%) |

**S5 Table:** Distribution and satisfaction rates for participants reporting “Other” treatment frequency across symptom burden subtypes (N=125). Free-test treatment frequency responses from participants selecting “other” in Table 3 were categorized into five clinically meaningful groups based on injection interval: loading phase, very frequent (every two weeks or less), extended standard (every 3-11 weeks, extended interval (every 12 weeks or more), and no treatment or pre-treatment. Satisfaction rates are presented as number satisfied/number responding (percentage), excluding participants who did not respond to the satisfaction question (High Burden: n=37; Moderate Burden: n=69; Low Burden: n=14 with Yes or No satisfaction response). Very frequent injections (every two weeks or less) were associated with the highest satisfaction rates across all subtypes reporting thus frequency (High Burden: 77・8%; Moderate Burden: 81・8%). No High Burden participants reported extended interval injections (every 12 weeks or more), while no High Burden participants with no established treatment reported satisfaction (0・0%, n=5). The single Moderate Burden participant with no treatment reporting satisfaction (100・0%) should be interpreted with caution given the small cell size. Loading phase satisfaction reflects treatment at an early stage where a maintenance frequency has not yet been established.

| Treatment Frequency | High Burden (n=39) |  | Moderate Burden (n=72) |  | Low Burden (n=14) |  |
| --- | --- | --- | --- | --- | --- | --- |
|  | n | n_satisfied (%) | n | n_satisfied (%) | n | n_satisfied (%) |
| Loading phase | 9 | 3/7 (42.9%) | 14 | 6/14 (42.9%) | NA | NA |
| Very frequent (<=2 weeks) | 9 | 7/9 (77.8%) | 12 | 9/11 (81.8%) | 1 | 1/1 (100.0%) |
| Extended standard (3-11 weeks) | 16 | 7/16 (43.8%) | 35 | 19/34 (55.9%) | 8 | 6/8 (75.0%) |
| Extended interval (>=12 weeks) | NA | NA | 9 | 5/9 (55.6%) | 5 | 4/5 (80.0%) |
| No treatment or pre-treatment | 5 | 0/5 (0.0%) | 2 | 1/1 (100.0%) | NA | NA |

## Supplementary Figures

**S1 Fig.**
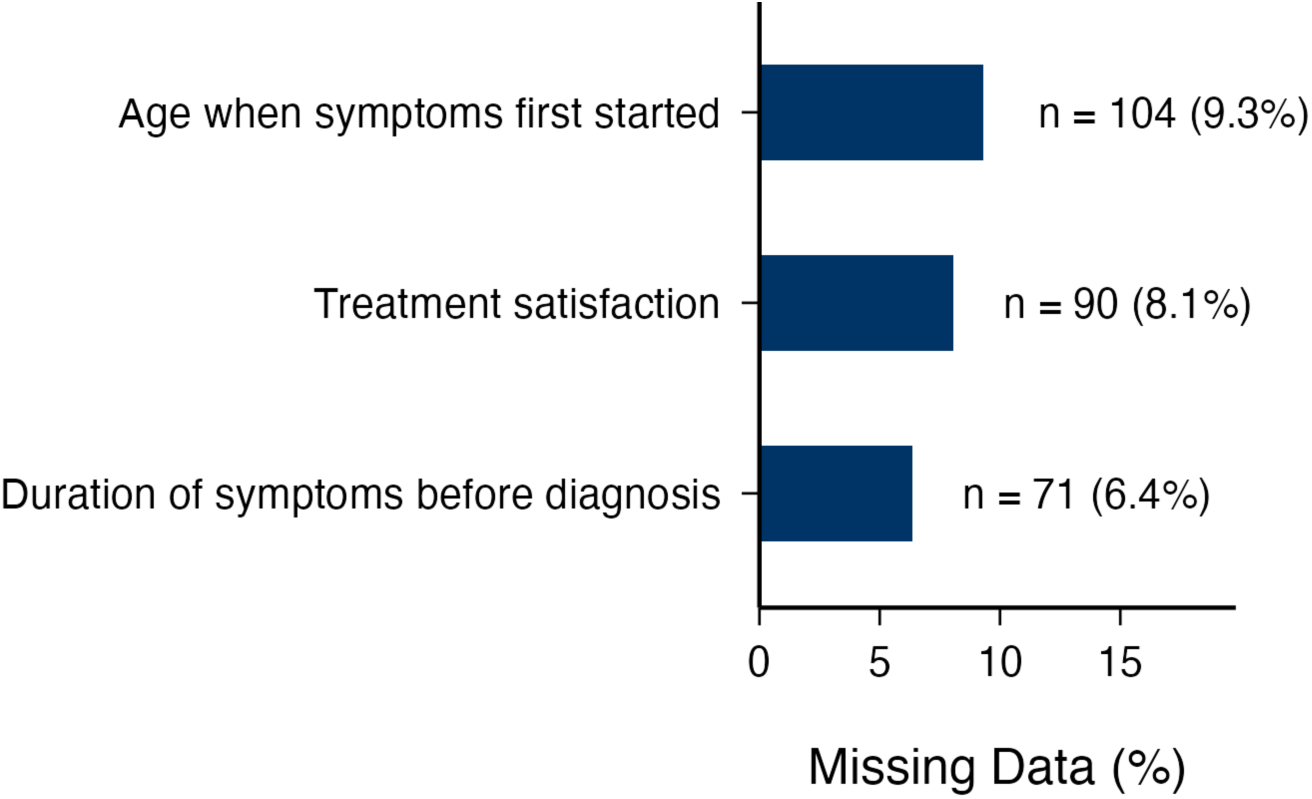
Missing data for post-hoc outcome variables. Percentage of missing values for post-hoc outcome variables in the analytic sample (N=1,117). Complete data were available for all 46 LCA indicator variables (symptoms, conditions, and family history variables) across the full analytic sample. Missing data were only observed in post-hoc outcome variables: age when symptoms first started (9·3%, n=104), duration of symptoms before diagnosis (6·4%, n=71), and treatment satisfaction (8·1%, n=90). Participants with missing post-hoc data were retained in the LCA but excluded listwise from the relevant post-hoc analysis.

**S2 Fig:**
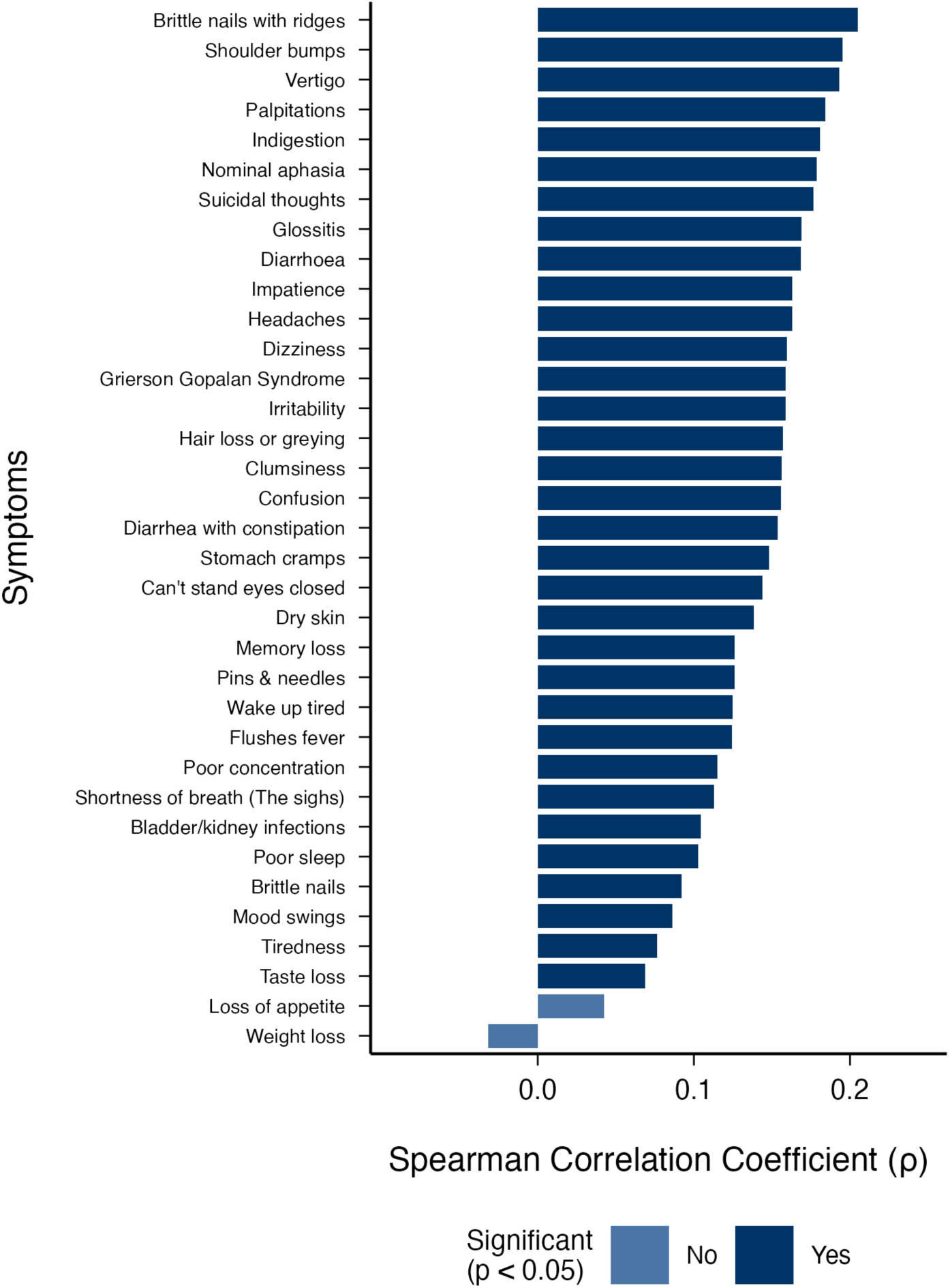
Spearman correlation coefficients. Spearman correlation coefficients between symptom duration before diagnosis and individual symptom endorsement for 35 symptoms (N=1,117). Dark blue bars represent statistically significant correlations (p<0·05), while light blue bars indicate non-significant correlations. While 33/35 symptoms (94·3%) showed statistically significant correlations, the magnitude was weak (range -0·032 to 0·205), suggesting minimal practical impact of recall bias on symptom reporting.

**S3 Fig:**
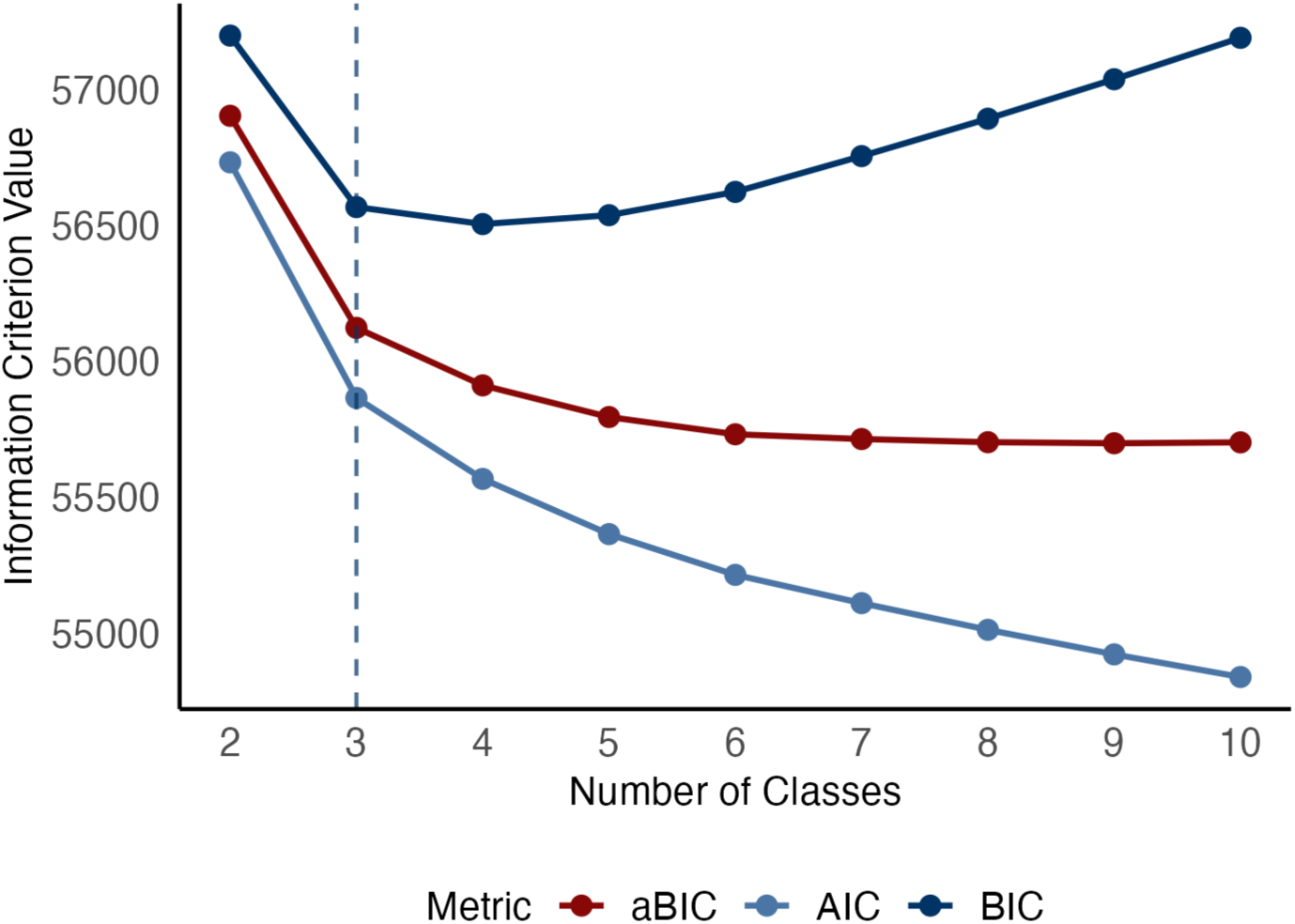
Model fit statistics for latent class analysis (N=1,117). Information criterion values (AIC, BIC, aBIC) for latent class models with 2-10 classes. Lower values indicate better model fit, with BIC penalizing model complexity more strongly than AIC and aBIC. The 3-class solution (indicated by dashed vertical line) was selected based on convergence of BIC and aBIC inflection points, a significant Lo-Mendell-Rubin test (p<0.0001), and adequate class size across all three groups. BIC showed substantial improvement from the 2-class (57200·10) to 3-class model (56568·60), with diminishing returns thereafter. While the 4-class model achieved a lower BIC value (56506·08), the Lo-Mendell-Rubin test indicated no significant improvement over the 3-class solution (p=0·51), supporting the 3-class solution as the optimal balance model fit and parsimony. AIC = Akaike Information Criterion; BIC = Bayesian Information Criterion; aBIC = sample-size adjusted BIC.

**S4 Fig:**
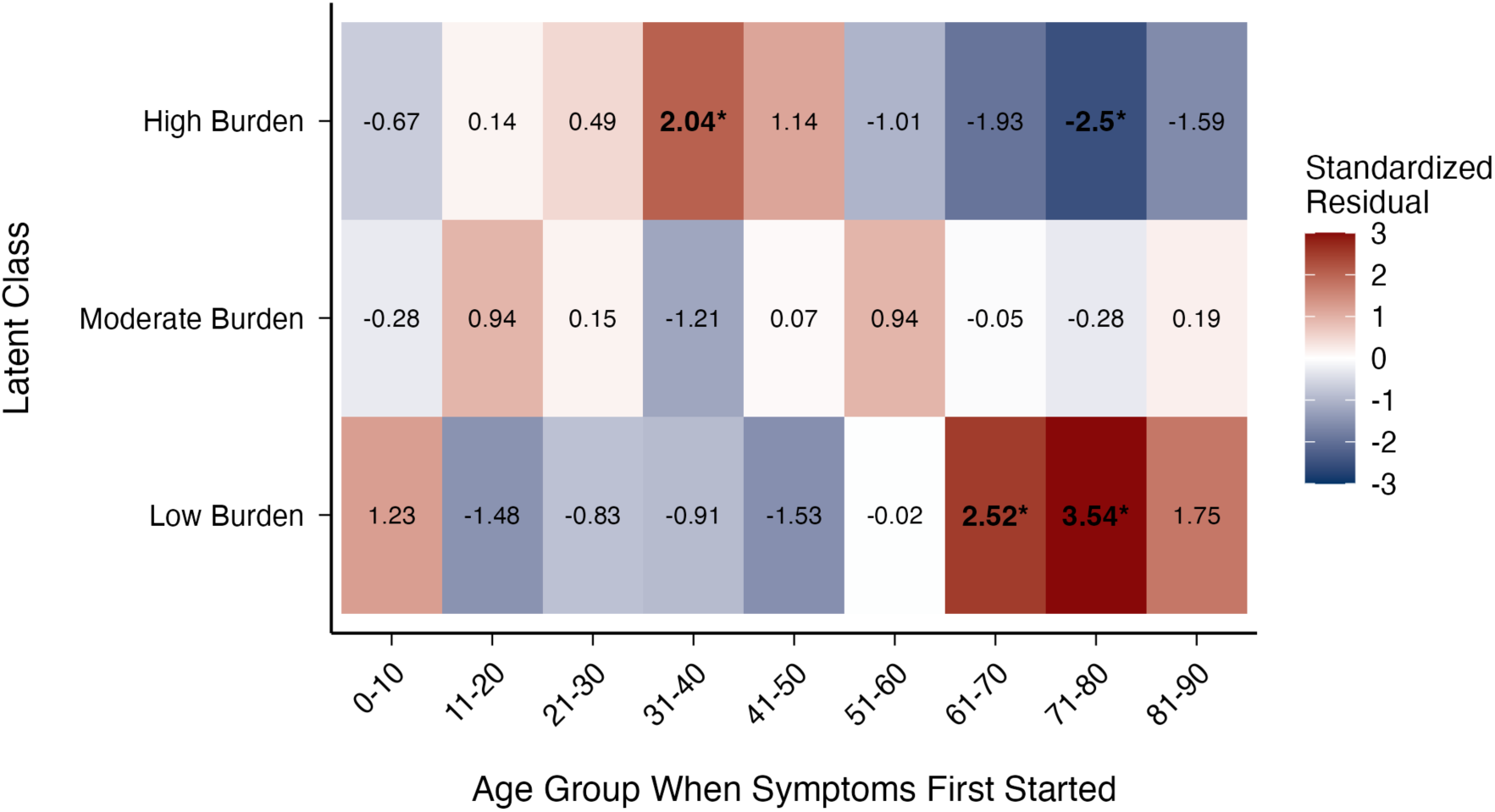
Standardized residuals from chi-squared analysis of age when symptom first started by symptom burden subtype (High Burden n=295, Moderate Burden n=563, Low Burden n=155, total n=1,013). Standardized residuals indicate the degree to which observed cell counts deviate from expected counts under independence. Positive values (red) indicate more observations than expected; negative values (blue) indicate fewer. Bold values with asterisks (*) denote statistically significant overrepresentations (|z|>1·96, p<0·05). The Low Burden subtype shows significant overrepresentation in the 61-70 (z=2·52) and 71-80 (z=3·54) age groups. The High Burden subtype shows significant overrepresentation in the 31-40 age group (z=2·04) and significant underrepresentation in the 71-80 age group (z=-2·50), suggesting that younger age of symptom onset is associated with higher symptom burden. Data were available for 1,013 of 1,117 participants (90·7%); participants with missing age data were excluded listwise.

**S5 Fig:**
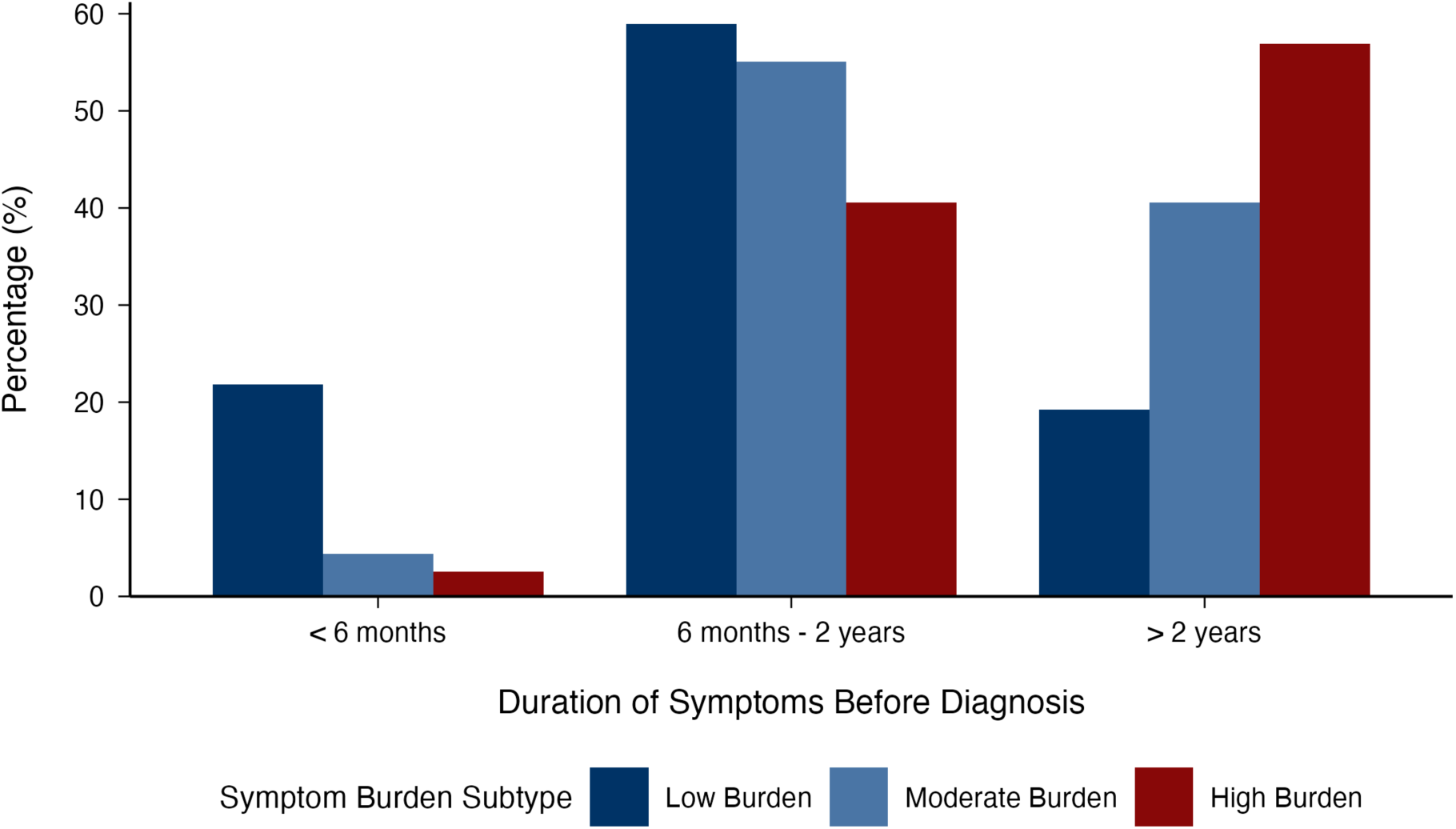
Duration of symptoms before diagnosis across symptom burden subtypes (High Burden n=318, Moderate Burden n=572, Low Burden n=156, total n=1,046). Values represent number and percentage of patients within each symptom burden subtype reporting each duration category. Duration categories are collapsed into three groups: less than 6 months, 6 months to 2 years, and more than 2 years (see **S4 Fig** for the full 11-category distribution). Chi-square analysis revealed highly significant differences in symptom duration before diagnosis (χ²=115·47, df=4, p<0·001, Cramér’s V=0·23), with High Burden subtype showing the longest pre-diagnosis periods compared to Moderate and Low Burden subtypes. Participants with missing data for symptom duration were excluded listwise (71 of 1,117 participants).

**S6 Fig:**
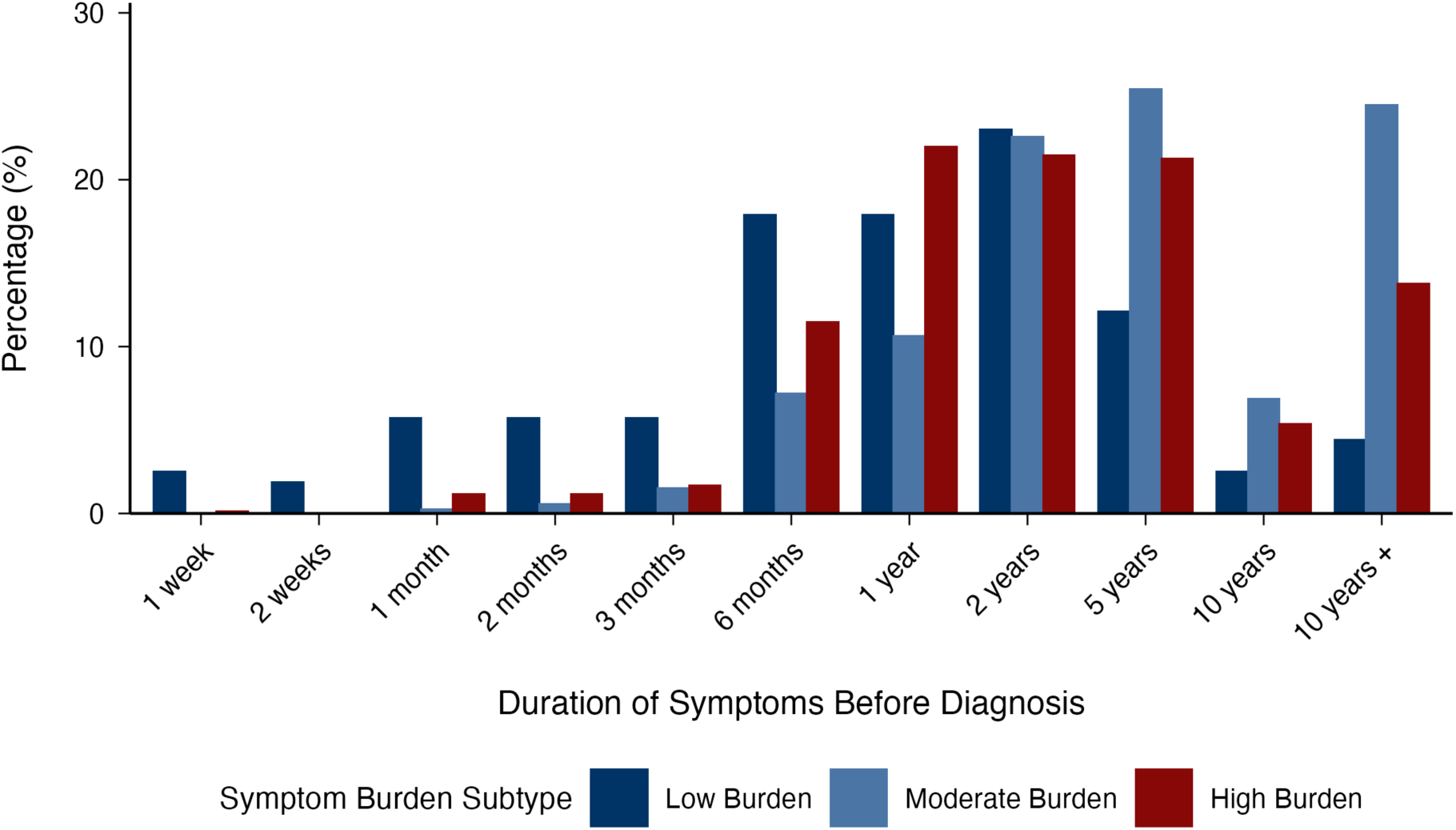
Duration of symptoms before diagnosis across symptom burden subtypes ∼ full distribution (High Burden n=318, Moderate Burden n=572, Low Burden n=156, total n=1,046). Percentage distribution of symptom duration before diagnosis across the three symptom burden subtypes, showing all 11 duration categories from 1 week to more than10 years. The High Burden subtype (dark red) shows the highest proportion of participants with prolonged symptom duration, particularly in the 5- year and 10+ year categories, while the Low Burden subtype (dark blue) demonstrates earlier diagnosis with higher proportions in shorter duration categories. Chi-square analysis confirmed significant association between symptom duration and class membership (χ²=149·59, df=20, p<0·001, Cramér’s V=0·27). Participants with missing data for symptom duration were excluded listwise (71 of 1,117 participants).

**Strengthening the Reporting of Observational Studies in Epidemiology (STROBE) Statement**

| Item<br>Title an Abstract | Item No | STROBE Guideline |
| --- | --- | --- |
|  | 1 | (a) Indicate the study's design with a commonly used term in the title or the abstract.<br>(b) Provide in the abstract an informative and balanced summary of what was done ad what was found. |
| **Introduction** |  |  |
| Background/rationale | 2 | Explain the scientific background and rationale for the investigation being reported. |
| Objectives | 3 | State specific objectives, including any prespecified hypotheses. |
| **Methods** |  |  |
| Study designs | 4 | Present key elements of study design early in the paper. |
| Setting | 5 | Describe the setting, locations, and relevant dates, including periods of recruitment, exposure, follow-up, and data collection. |
| Participants | 6 | *Cross-sectional study*-Give the eligibility criteria, and the sources and methods of selection of participants. |
| Variables | 7 | Clearly define all outcomes, exposures, predictors, potential confounders, and effect modifiers. Give diagnostic criteria, if applicable. |
| Data sources/measurements | 8 | For each variable of interest, give sources of data and details of methods of assessment (measurement). Describe comparability of assessment methods if there is more than one group. |
| Bias | 9 | Describe any efforts to address potential sources of bias. |
| Study size | 10 | Explain how the study size was arrived at. |
| Quantitative variables | 11 | Explain how quantitative variables were handled in the analysis. If applicable, describe which groupings were chosen and why. |
| Statistical methods | 12 | (a) Describe all statistical methods, including those used to control for confounding.<br>(b) Describe any methods used to examine subgroups and interactions.<br>(c) Explain how missing data were addressed.<br>(d) *Cross-sectional study* - If applicable, describe analytical methods taking account of sampling strategy.<br>(e) Describe any sensitivity analysis. |
| **Results** |  |  |
| Participants | 13 | (a) Report numbers of individuals at each stage - e.g., numbers potentially eligible, examined for eligibility, confirmed eligibility, included in the study, completing follow-up, and analyzed.<br>(b) Give reasons for non-participation at each stage.<br>(c) Consider use of a flow diagram. |
| Descriptive data | 14 | (a) Give characteristics of study participants (e.g., demographic, clinical, social,) and information on exposures and potential confounders.<br>(b) Indicate number of participants with missing data for each variable of interest. |
| Outcome data | 15 | *Cross-sectional study* - Report numbers of outcome events or summary measures. |
| Main results | 16 | (a) Give unadjusted estimates and, if applicable, confounder-adjusted estimates and their precision (e.g., 95% confidence interval). Make clear which confounders were adjusted for and why they were included.<br>(b) Report category boundaries when continuous variables were categorized.<br>(c) If relevant, consider translating estimates of relative risk into absolute risk for a meaningful time period. |
| Other analysis | 17 | Report other analysis done - e.g., analysis of subgroups and interactions and sensitivity analyses. |
| **Discussion** |  |  |
| Key results | 18 | Summarize key results with reference to study objectives. |
| Limitations | 19 | Discuss limitations of the study, taking into account sources of potential bias or imprecision. Discuss both direction and magnitude of any potential bias. |
| Interpretation | 20 | Give a cautious overall interpretation of results considering objectives, limitations, multiplicity of analyses, results from similar studies, and other relevant evidence. |
| Generalizability | 21 | Discuss the generalizability (external validity) of study results. |
| **Other information** |  |  |
| Funding | 22 | Give the source of funding and the role of the funders for the present study and, if applicable, for the original study on which the present article is based. |

## Notes

### Competing Interest Statement

Katrina Burchell is CEO of the Pernicious Anaemia Society (unpaid) and Chair of the B12 Alliance (CLuB12, unpaid). Andrew McCaddon receives royalty payments from COBALZ Limited relating to a medical food (Cerefolin NAC) licensed to Alfa-Sigma. Marie-Joe Dib declares grant support from Nascent Studio Ltd, UK. Austin W. Reynolds declares grant support from the National Science Foundation, USA. All other authors declare no competing interests.

### Author Declarations

The NHS Research Ethics Committee (NRES) waived ethical approval for this work. The original data collection was classified as a service evaluation audit rather than clinical research, as it was designed solely to evaluate current care and did not involve randomization or allocation to any intervention (NRES, 2009).

